# Human Mobility Restrictions and the Spread of the Novel Coronavirus (2019-nCoV) in China

**DOI:** 10.1101/2020.03.24.20042424

**Authors:** Hanming Fang, Long Wang, Yang Yang

## Abstract

We quantify the causal impact of human mobility restrictions, particularly the lockdown of the city of Wuhan on January 23, 2020, on the containment and delay of the spread of the Novel Coronavirus (2019-nCoV). We employ a set of difference-in-differences (DID) estimations to disentangle the lockdown effect on human mobility reductions from other confounding effects including panic effect, virus effect, and the Spring Festival effect. We find that the lockdown of Wuhan reduced inflow into Wuhan by 76.64%, outflows from Wuhan by 56.35%, and within-Wuhan movements by 54.15%. We also estimate the dynamic effects of up to 22 lagged population inflows from Wuhan and other Hubei cities, the epicenter of the 2019-nCoV outbreak, on the destination cities’ new infection cases. We find, using simulations with these estimates, that the lockdown of the city of Wuhan on January 23, 2020 contributed significantly to reducing the total infection cases outside of Wuhan, even with the social distancing measures later imposed by other cities. We find that the COVID-19 cases would be 64.81% higher in the 347 Chinese cities outside Hubei province, and 52.64% higher in the 16 non-Wuhan cities inside Hubei, in the counterfactual world in which the city of Wuhan were not locked down from January 23, 2020. We also find that there were substantial undocumented infection cases in the early days of the 2019-nCoV outbreak in Wuhan and other cities of Hubei province, but over time, the gap between the officially reported cases and our estimated “actual” cases narrows significantly. We also find evidence that enhanced social distancing policies in the 63 Chinese cities outside Hubei province are effective in reducing the impact of population inflows from the epicenter cities in Hubei province on the spread of 2019-nCoV virus in the destination cities elsewhere.

**JEL Codes:** I18, I10.

## 1 Introduction

Human mobility contributes to the transmission of infectious diseases that pose serious threats to global health. Indeed, in response to pandemic threats many countries consider and impose measures that restrict human mobility flows as one of their response plans (Bajardi et al., 2011; Wang and Taylor, 2016; Charu et al., 2017). However, restrictions on human mobility are controversial not only because of their negative economic impacts, but also because of the uncertainty about their effectiveness in controlling the epidemic. Even if restricting human movement could lead to improvements in disease control and reductions in health risks, it is empirically challenging to quantify the impact of human mobility on the spread of infectious diseases, and to understand the detailed spatial patterns of how the infectious disease spreads. Both granular disease occurrence data and human mobility data (Charu et al., 2017) are hard to obtain; moreover, it is difficult to disentangle the impact of human mobility from other potential contributing factors in the spread of epidemics (Ferguson et al., 2006; Hollingsworth et al., 2006). In this paper, we exploit the exogenous variations in human mobility created by lockdowns of Chinese cities during the outbreak of the Novel Coronavirus (2019-nCoV), and utilize a variety of high-quality data sets, to study the effectiveness of an unprecedented *cordon sanitaire* of the epicenter of COVID-19, and provide a comprehensive analysis on the role of human mobility restrictions in the delaying and the halting of the spread of the COVID-19 pandemic.^1^

The fast-moving 2019-nCoV that infected 266,073 people and claimed 11,184 lives as of March 21, 2020 is deteriorating into one of the worst global pandemics.^2^ The virus emerged in the city of Wuhan in the Hubei Province of China in early December of 2019, spread mainly through person-to-person contact (Chan et al., 2020), and rapidly reached more than 183 countries as of March 21, 2020.^3^ Currently, there are no licensed vaccines or specific therapeutics to combat COVID-19. The lockdown of 11 million people in Wuhan from January 23, 2020 represents by then the largest quarantine in public health history, and offers us an opportunity to rigorously examine the effects of the city lockdown and understand the relationship between human mobility and virus transmission.

Specifically, this paper studies five research questions. First, how does the lockdown of the city of Wuhan amid the Novel Coronavirus outbreak affect population movement? Second, how do population flows among Chinese cities, particularly outflows from Wuhan and other cities in Hubei province, affect virus infection in the destination cities? Third, is there evidence of, and if so, what is the magnitude of, undocumented cases of COVID-19 cases in Wuhan and other cities in Hubei province during the early stages of the epidemic? And how does the extent of undocumented infection cases evolve over time? Fourth, how many COVID-19 cases elsewhere in China were prevented by the unprecedented Wuhan lockdown? Fifth, are social distancing policies in destination cities effective in reducing the spread of the infections? We utilize reliable datasets on population migration among pairs of Chinese cities and the within-city population movements of each city at the daily level from Baidu Migration, and the city-level daily numbers of confirmed COVID-19 cases, recovered patients, and death tolls from the Chinese Center for Disease Control and Prevention (CCDC) during a sample period of January 1-February 29, 2020, covering 22 days before and 38 days after the city lockdown on January 23, 2020, as well as the matched data from the same lunar calendar period in 2019.

We first employ various difference-in-differences (DID) estimation strategies to disentangle the effect of Wuhan lockdown on human mobility reductions from other confounding effects including panic effect, virus effect, and the Spring Festival effect–the Spring festival of the Chinese New Year is on January 25, 2020 (Table 2). We find that the lockdown of Wuhan reduced inflow into Wuhan by 76.64%, outflows from Wuhan by 56.35%, and within-Wuhan movements by 54.15%. We also estimate the dynamic effects of up to 22 lagged population inflows from Wuhan and other Hubei cities, the epicenter of the 2019-nCoV outbreak, on the destination cities’ new infection cases (Figure 4). We discover that the estimated effects of the different lags of inflows from Wuhan and Hubei both show a clear inverted *U* -shape with respect to the lags, with the largest impact on the newly confirm cases today comes from the inflow population from Wuhan or other cities in Hubei about 12 to 14 days earlier. We find, using simulations with these estimates, that the lockdown of the city of Wuhan on January 23, 2020 contributed significantly to reducing the total infection cases outside of Wuhan, even with the social distancing measures later imposed by other cities. We find that the COVID-19 cases would be 64.81% higher in the 347 Chinese cities outside Hubei province, and 52.64% higher in 16 non-Wuhan cities inside Hubei, in the counterfactual world in which the city of Wuhan were not locked down from January 23, 2020. We also find that there were substantial undocumented infection cases in the early days of the 2019-nCoV outbreak in Wuhan and other cities of Hubei province, but over time, the gap between the officially reported cases and our estimated “actual” cases narrows significantly. We also find evidence that imposing enhanced social distancing policies in the 63 Chinese cities outside Hubei province is effective in reducing the impact of population inflows from the epicenter cities in Hubei province on the spread of 2019-nCoV virus in the destination cities elsewhere.

By providing a rigorous estimation of the impact of within and cross-city migration on the spread of the 2019-nCoV virus in China, our study contributes to fast-growing literature on 2019-nCoV infection, mostly in the medical and public health fields. Huang et al. (2020) describe high rates of respiratory distress, intensive care admission, and abnormal findings on chest computed tomography (CT) in the first 41 patients hospitalized from December 16 to January 2 in Wuhan, as well as a 15% death rate. Chan et al. (2020) investigate a family cluster and confirm the human-to-human transmission of this Novel Coronavirus within hospitals and families. The basic reproduction number (R0) for COVID-19 in 12 studies has a mean of 3.28 and a median of 2.79 (Liu et al., 2020) (compared to 3 for SARS (WTO, 2003)). This study is also related to disaster-induced migration, which has often occurred during flooding (Gray and Mueller, 2012), drought (Munshi, 2003), earthquake (Lu et al., 2012), and other destructive climatic phenomena.

Given that the 2019-nCoV has rapidly spread worldwide due to human travel and caused severe illness and significant mortality, it is therefore essential to understand the impact of various control measures on human mobility and the virus transmission. Qiu et al. (2020) apply machine learning tools and use exogenous temperature, wind speed, and precipitation in the preceding third and fourth weeks as the instruments to show that the massive lockdown and other control measures significantly reduced the virus transmission. Their results highlight that the population outflow from the outbreak source city poses higher risks to the destination cities than other social and economic factors, such as geographic proximity and similarity in economic conditions. Our results are also in line with the results of the latest modeling exercises, which mostly rely on model calibrations of various parameters, such as generation time, incubation period, detection rates, and changes in travel flow. Using the Global Epidemic and Mobility model, Chinazzi et al. (2020) project the impact of travel limitation on the national and international spread of the 2019-nCoV. The study finds that only 24.4% of infected cases were reported as of February 1, and the Wuhan lockdown reduced the cases by 10% in cities outside Wuhan by January 31. The study most closely related to ours is Li et al. (2020), which shows that contagious but undocumented COVID-19 cases facilitated the geographic spread of the epidemic in China. Using a networked dynamic meta-population model and Bayesian inference, Li et al. (2020) find that 86% of all infections were undocumented before the Wuhan lockdown and reported infections would have been reduce by 78.7% in China, without transmission from undocumented cases between January 10 and January 23. Lai et al. (2020) build a travel network-based susceptible-exposed-infectious-removed (SEIR) model to simulate the outbreak across cities in mainland China. They use epidemiological parameters estimated from the early stage of outbreak in Wuhan to parameterize the transmission before the non-pharmaceutical interventions (NPI) were implemented. The NPIs they consider include travel bans and restrictions, contact reductions and social distancing, early case identification and isolation. Through their sim-ulations, they find that the NPIs deployed in China appear to be effectively containing the COVID-19 outbreak, but the efficacy of the different interventions varied, with the early case detection and contact reduction being the most effective. Moreover, deploying the NPIs early is also important to prevent further spread. Relative to our study, it is important to point out that in their simulations, they assumed that the pattern of population movements was the same in years when there were no outbreaks and interventions. To the best of our knowledge, this paper is the first to provide a *causal* interpretation of the impact of city lockdown on human mobility and the spread of 2019-nCoV, and to clearly disentangle the lockdown effects from other potential contributing factors such as panic and virus effect, as well as the seasonal Spring Festival effect (see Section 3).

The remainder of the paper is structured as follows. In Section 2, we describe the data sets used in our analysis. In Section 3, we present different difference-in-difference estimation strategies to separate the lockdown effect, panic effect, virus (deterrence) effect, and the Spring Festival effect on population movements in China. In Section 4, we estimate the distributed lag effects of inflows from the epicenter cities of the 2019-nCoV outbreak on destination cities’ daily infection cases. In Section 5, we study how the enhanced social distancing policies, or “lockdown” policies, in the destination cities’ impact the effects of the population inflows from the epicenter cities. In Section 6, we conclude.

## 2 Data and Descriptive Statistics

### Population Migration Data

We obtain inter-city population migration data from Baidu Migration, a travel map offered by the largest Chinese search engine, Baidu.^4^. The Baidu Migration data set covers 120,142 pairs of cities per day for 364 Chinese cities between January 12 and March 12 in 2019, and between January 1 and February 29 in 2020. Note that, by the lunar calendar, the data covers the same period of 24 days before and 36 days after the Spring Festivals, respectively for year 2019 and year 2020. The daily inter-city migration data consist of 2,977,899 city-pair observations each year. In addition, Baidu provides the daily *within-city* mobility data for each city in the sample period, which is a panel consisting of 21,655 city-day level observations each year.

The Baidu Migration data is based on real-time location records for every smart phone using the company’s mapping app, and thus can precisely reflect the population movements between cities.^5^ For each of the 364 cities, Baidu Migration provides the following informa-tion: (1). the top 100 origination cities (OC) for the population moving *into* the city and the corresponding percentages of inflow population that originated from each of the top 100 OC; (2). the top 100 destination cities (DC) for the population moving *out of* the city, and the corresponding percentages of the outflow population that go into each of the top DC. In the data, the cumulative percentages of the inflow population from the top 100 origination city, and the cumulative percentages of the outflow population into the top 100 destination cities, reach 97% per city on average. This ensures that the Baidu Migration data capture near complete inflows and outflows for each of the 364 cities in the data.

In addition, the Baidu Migration data provides three migration intensity indicators: the daily *in-migration* index (IMI) of a city, the daily *out-migration* index (OMI) of a city, and the daily *within-city* migration index (WCMI). The intensity indicators are consistent across cities and across time. To convert the index to the number of people, we use the actual number of inflow population into Shanghai by airplanes, trains and buses/cars, and the number of within-Shanghai trips using subways, buses and expressways, collected by the National Earth System Science Data Center for the period of February 6 to February 22, 2020. Using this data, we estimate that one index unit in the IMI and OMI corresponds to 90,848 person movements, and one index unit in WCMI corresponds to 2,182,264 person movements. We are thus able to calculate the *number* of daily inflow and outflow migrants in each city-pair, as well as the *number* of within-city population movements.^6^

Table 1 presents the summary statistics of the population flows at the city-pair-day level and city-day level. It shows drastic declines in the average inflows, average outflows, and average within-city migration in 2020, compared to a sample period in 2019 (matched by the lunar calendar date). The plummeting of the migration statistics due to the Wuhan lockdown is also depicted in Figure 2. The three figures on the top show the inflows into Wuhan, outflows from Wuhan and within-Wuhan flows, for year 2020 (solid line) and year 2019 (dashed line), matched by the lunar calendar; and the bottom shows the corresponding figures for the national city averages. The first vertical line indicates the date of January 20, 2020 when experts confirmed that 2019-nCoV could transmit from human to human; and the second vertical line indicates the date of January 23, 2020 when Wuhan was locked down. It is clear that while the flows in 2020 tracked that of 2019 well until January 20, the 2020 level dropped to a fraction of their corresponding 2019 levels, particularly after the lockdown of Wuhan.

**Table 1:** Descriptive Statistics

| **Panel A: City-Pair-daily Level Data** |  |  |  |  |
| --- | --- | --- | --- | --- |
| Sample Period | Jan 12, 2019 - Mar 12 , 2019 |  | Jan 1, 2020 - Feb 29, 2020 |  |
|  | Mean | S.D | Mean | S.D |
| CityPair Intensity | 0.011 | 0.052 | 0.007 | 0.043 |
| CityPair Flow Population | 989.849 | 4,692.64 | 665.766 | 3,944.00 |
| **Panel B: City-daily Level Data** |  |  |  |  |
| Sample Period | Jan 12, 2019 - Mar 12 , 2019 |  | Jan 1, 2020 - Feb 29, 2020 |  |
|  | Mean | S.D | Mean | S.D |
| Outflow Intensity | 1.168 | 1.572 | 0.723 | 1.473 |
| Outflow Population | 106,153.47 | 142,836.35 | 65,659.22 | 133,811.95 |
| Inflow Intensity | 1.164 | 1.599 | 0.720 | 1.078 |
| Inflow Population | 105,703.22 | 145,263.72 | 65,451.07 | 97,892.32 |
| WithinCity Flow Intensity | 4.475 | 0.814 | 3.606 | 1.514 |
| WithinCity Flow Population | 1,502,229.73 | 1,348,525.551 | 1,198,683.924 | 1,226,171.714 |
| # of New Confirmed Cases | 0 | 0 | 3.559 | 104.18 |
| # of Total Confirmed Cases | 0 | 0 | 75.973 | 1,317.66 |
| # of New Deaths | 0 | 0 | 0.128 | 3.017 |
| # of Total Deaths | 0 | 0 | 2.156 | 51.414 |
| # of New Heals | 0 | 0 | 1.819 | 34.077 |
| # of Total Heals | 0 | 0 | 17.623 | 288.538 |

*Notes:* This table presents the descriptive statistics of the variables used in this study, with Panel A corresponding to the city-pair daily level variables and Panel B corresponding to the city daily level variables. The sample covers a period between Jan 12 and Mar 12 in 2019, and between Jan 1 and Feb 29 in 2020. The two periods in 2019 and 2020 cover the same lunar calendar.

**Table 2:**
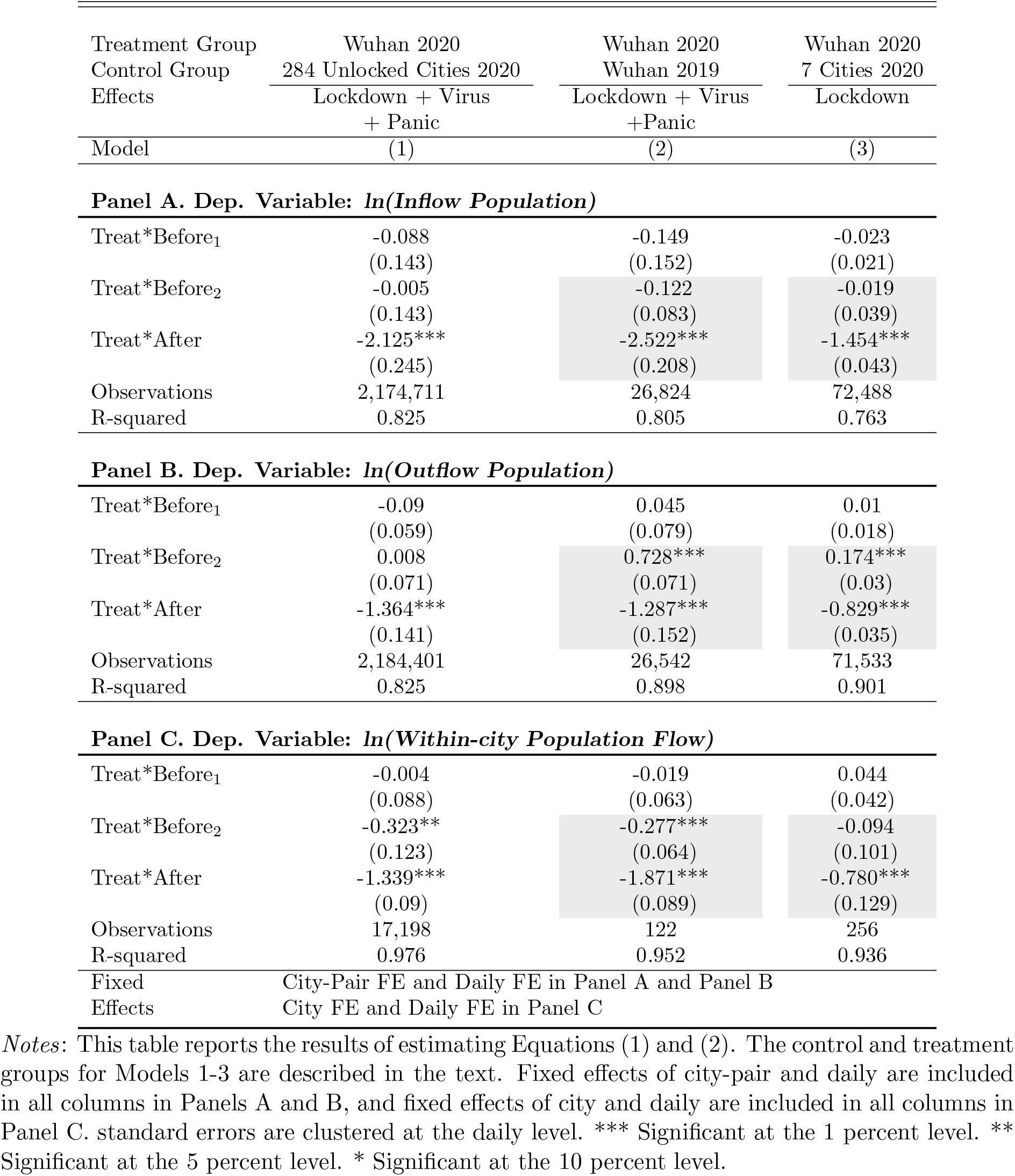
The Effects of Wuhan Lockdown on Population Movement

### Outbreak Data

COVID-19 daily case counts are collected from China CDC, which provides daily updates on confirmed, dead, and recovered COVID-19 cases in each city.^7^ From January 11 to February 29, 2020, the data covers 77,105 laboratory-confirmed COVID-19 cases, 2,778 death cases, and 39,392 recovered cases in 296 cities in China. Panel B of Table 1 presents the summary statistics of COVID-19 data, and Figure 3 plots the trends of daily statistics of COVID-19separately for the epicenter city of Wuhan, for other cities in Hubei Province, and for cities outside of Hubei.^8^

There are many possible reasons for us to treat the officially reported numbers of confirmed cases in Wuhan and other cities of Hubei with more caution, and to treat them differently from the data of cities outside of Hubei. As the epicenter of COVID-19, the health care systems in Wuhan and other cities in Hubei were overwhelmed by the sheer number of patients who needed laboratory testing, especially in the early phases of the virus outbreak. As such, the over-extended medical system in Wuhan and other cities in Hubei might have caused delays in the testing of the patients who contracted COVID-19; and because of the delay, some patients who contracted COVID-19 might have self-recovered, or might have died, before being officially tested; and some who were infected with the virus might be asymptomatic. There is also a possibility that government officials in the epicenter cities had incentives to downplay the severity of the outbreak, at least initially. These considerations impact how we use the outbreak data in Section 4.

## 3 The Impact of Wuhan Lockdown on Population Movements

The incubation period of 2019-nCoV is long in comparison with SARS; moreover, the virus can transmit while the person is still asymptomatic, which increases the probability a person with the Novel Coronavirus will travel and unknowingly spread the virus to others.^9^ To suppress the spread of 2019-nCoV, the central government of China imposed an unprecedented lockdown in Wuhan starting from 10 am of January 23, 2020, and in other Hubei cities several days later. As of February 29, 2020, 80 cities in 22 provinces issued different levels of lockdown policies.^10^ Table A1 in the Appendix provides the detailed information about the various forms of population mobility control in different cities. We also plot the geographic distributions of sample cities and cases in Figure 1.

**Figure 1:**
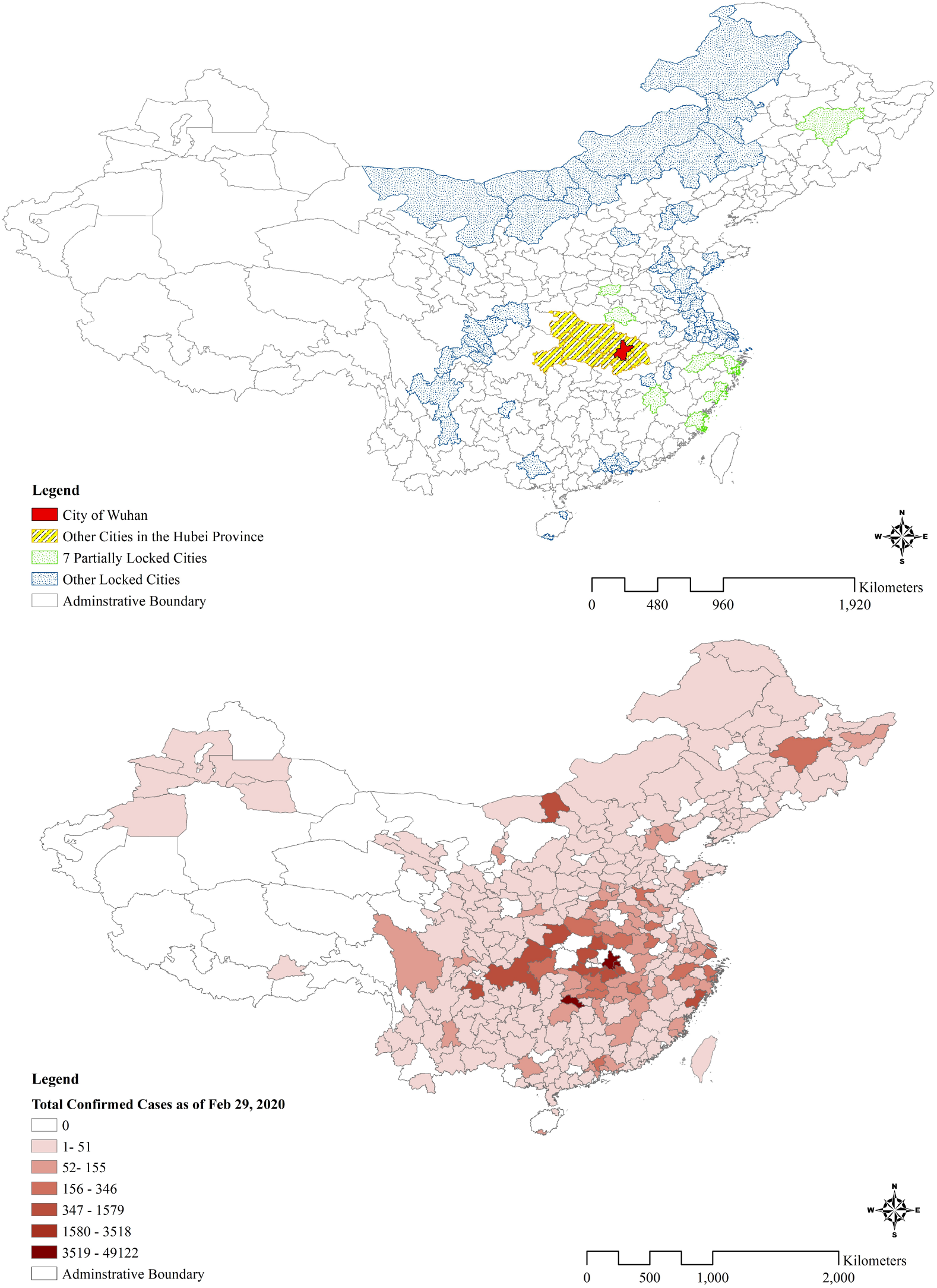
Cities with Control Measures and the Confirmed COVID-19 Cases. *Notes*: This figure presents the geographic distributions of cities with different levels of control measures and the number of Confirmed COVID-19 Cases as of Feb 29, 2020. The maps were plotted with ArcGIS 10.2 (ESRI).

### 3.1 Empirical Challenges

There are several confounding factors in our attempt to *causally* quantify the impact of lockdown on human mobility, and on the spread of infectious viruses. First, the virus outbreak happens right before the Spring Festival of the Chinese Lunar New Year, which causes the largest annual human migration every year.^11^ Second, the virus itself, even *in the absence* of a mandatory lockdown, may lead to curtailed human movement as people attempt to avoid exposure to the virus in the journeys and public spaces. We refer to this deterrence effect as the *virus effect*. Third, for the city of Wuhan and other cities in Hubei that are close to Wuhan, there is also the possibility of a *panic effect*, in reaction to the virus. The panic effect can lead to an increase in the population outflow from the epicenter of the virus outbreak, and a decrease of the population inflow to the epicenter, particularly the city of Wuhan. The panic effect is likely to peak when the government officially confirmed on January 20, 2020 that the Novel Coronavirus can transmit from person-to-person. In our analysis below, we create a specific pre-lockdown period *Before*_2,*t*_, which includes the three-day period between January 20 and January 22, 202, to capture the panic effects.^12^ Note that while the virus effect applies to movement into and out of all cities, the panic effect is more specific to the cities in the epicenter, especially Wuhan, and can have a positive effect on outflows, and a negative effect on inflows.

### 3.2 Effects of Various Factors on Inter-City Population Mobility

We first examine the impact of city lockdown on inter-city population mobility, including inflow and outflow, between a city pair (*i, j*). To disentangle the contributions of these confounding factors on human mobility, we exploit many unique sources of variations in the data, and employ several DID estimation strategies by comparing different treatment and control groups. The DID specification can be described as follows:

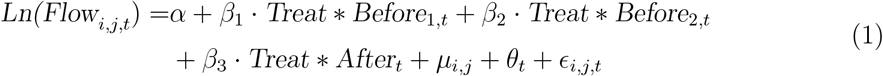

where *i, j*, and *t* respectively index the destination city, origination city, and date; the dependent variable, *Ln(Flow*_*i,j,t*_), is the logarithmic population flows received by city *i* from city *j* at date *t*. The definition of *Treat* varies by specific DID designs, and we will be explicit about its definition below. The city-pair fixed effect *µ*_*i,j*_ is included to absorb the city-specific and the city-pair specific heterogeneities that may contaminate the estimation of our interested coefficient *β*_3_. We also control for the date-fixed effect *θ*_*t*_ to eliminate the time-specific impact, including the Spring Festival travel effect. The standard errors are clustered at the daily level.

In Equation (1), we include two pre-lockdown period indicators: *Before*_1,*t*_ is a dummy that takes value 1 for the period from January 11 to January 19, 2020 (4 to 11 days before the Wuhan lockdown), which can be used to examine the parallel trend assumption in the DID analysis; *Before*_2,*t*_ is a dummy that takes value 1 for the period from January 20 to January 22, 2020, three days before the unprecedented Wuhan lockdown, but after the official announcement that the Novel Coronavirus can spread from person to person.^13^ *Before*_2,*t*_ allows us to capture the panic effect. Finally, *After*_*t*_ is a dummy that takes value 1 for the sample period after the Wuhan lockdown, between January 23 and February 29, 2020. The omitted benchmark period is from January 1 to January 10, 2020.

### 3.3 Effects of Various Factors on Within-City Population Mobility

We also estimate the effect of lockdown on the within-city population movement utilizing the city-level data and a variety of DID specifications:

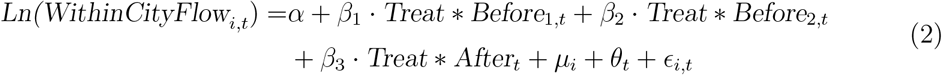

where *i* and *t* index the city and date. *Ln(WithinCityFlow*_*i,t*_) is the logarithmic within-city population mobility measure for city *i* at date *t*. Similar to Equation (1), *Treat* will be defined according to the DID design. *Before*_1,*t*_, *Before*_2,*t*_ and *After*_*t*_ are defined in the same way as in Equation (1). We include the city fixed effects *µ*_*i*_ and date fixed effects *θ*_*t*_. The standard errors are clustered at the daily level.

### 3.4 Estimation Results

In Table 2, we report the results from three sets of regressions specified according to Equation (1) for inter-city inflows (Panel A) and outflows (Panel B), and according to Equation (2) for within-city movement (Panel C). We implement three models that differ in the estimation sample, and the definition of the variable *Treat*.

#### Model 1: Wuhan 2020 vs. 284 Unlocked Cities in 2020

The estimation sample used in the regressions reported in Column (1) is the 2020 data for Wuhan and 284 cities that were never locked down during the Coronavirus outbreak.^14^

In Panel A when we examine the inflow population, *Treat* takes value 1 if the destination city *i* is Wuhan; in Panel B when we examine the outflow population, *Treat* takes value 1 if the origination city *j* is Wuhan. The control group consists of 284 cities that are not subject to any mobility restrictions. In Panel C when we examine the within-city mobility, *Treat* takes value 1 if city *i* is Wuhan. We interpret the coefficient estimate of *Treat* * *Before*_2,*t*_as measuring the panic effect of Wuhan relative to the unlocked cities; and the coefficient estimate of *Treat* * *After*_*t*_ as measuring the lockdown effect.^15^ The coefficient estimate of *Treat* * *Before*_1,*t*_ allows us to examine whether the parallel trend assumption for DID is satisfied. It is important to note that the possibly time-varying Spring Festival effects and the virus effects are both absorbed in the day fixed effects.

Based on the coefficient estimate of the term *Treat* * *After*_*t*_, we find that Wuhan’s lockdown reduces the inflow population *to* Wuhan by 88.06% (= 1 − *exp*(−2.215)), the outflow population *from* Wuhan by 74.44% (= 1 − *exp*(−1.364)), and within-city population movements in Wuhan by 73.79%(= 1 − *exp*(−1.339)), relative to all other unlocked cities in the post-lockdown period in 2020. In Panel C, we also find that the coefficient of *Treat*\**Before*_2,*t*_ is significantly negative at −0.323, suggesting that the official confirmation of the person-to-person transmission reduces the within-city movement in Wuhan by 27.6% from Jan 20 to Jan 22 in 2020. This points to a panic effect for the within-city population flow in Wuhan, but we do not observe a significant panic effect for inter-city flows in and out of Wuhan in Model 1.

#### Model 2: Wuhan 2020 vs. Wuhan 2019

Model 1 could be criticized on the ground that Wuhan may not be comparable to the 284 unlocked cities – after all, these 284 cities that never imposed any mobility restrictions in the COVID-19 outbreak could be very different from Wuhan. In Model 2, we compare the population movements of Wuhan in 2020 to *itself* in the same matched lunar calendar period in 2019, during which Wuhan is free of lockdown and Coronavirus outbreak. Thus, the estimation sample in Model 2 is the daily inflows into and outflows out of Wuhan, as well as the daily within-city movements in Wuhan for years 2019 and 2020.

Under Model 2, in Panel A when we examine the inflow population, *Treat* takes value 1 if the destination city *i* is Wuhan and year is 2020; in Panel B when we examine the outflow population, *Treat* takes value 1 if the origination city *j* is Wuhan and the year is 2020. The control group is Wuhan 2019. In Panel C when we examine the within-city mobility, *Treat* takes value 1 if the year is 2020. Under Model 2, the coefficient estimate of *Treat* * *Before*_2,*t*_ measures the panic effect related to the virus outbreak in Wuhan; and the estimate of *Treat* * *After*_*t*_ measures the sum of the *lockdown* effect and the *virus* effect. The coefficient estimate of *Treat* * *Before*_1,*t*_ still allows us to examine whether the parallel trend assumption for DID is satisfied. The day fixed effects absorb the possibly time-varying Spring Festival effects. Notice that the interpretation of the coefficient estimate of the term *Treat* * *After*_*t*_ differs from that in Model 1 because of the differences in the treatment and control groups.

Under Model 2, the estimated coefficients on *Treat* * *After*_*t*_, which, as we explained above, capture *both the lockdown and the virus effects*, remain negative, and economically and statistically significant in all panels. The estimates suggest that the lockdown of Wuhan, together with the deterrence effect of the virus (the virus effect), on average reduces the inflow population into, outflow population from, and within-city movements in Wuhan by 91.97% (= 1 − *exp*(−2.522)), 72.39% (= 1 − *exp*(−1.287)), and 69.49% (= 1 − *exp*(−1.871)), respectively, relative to the same lunar calendar days in 2019. We also find that the coefficient on *Treat* * *Before*_2,*t*_ is significantly positive in Panel B and significantly negative in Panel C, suggesting that the official confirmation of person-to-person spread of COVID-19 creates a panic effect, causing an increase of outflow from Wuhan of 107.09% (= *exp*(0.728) − 1), and a decrease of within-city movements in Wuhan of 24.19% (= 1 − *exp*(−0.277)), during the three days after the announcement but before the city lockdown. However, we do not observe a statistically significant panic effect for the population inflow into Wuhan, suggesting that people in other cities were not yet sufficiently concerned about the virus outbreak in Wuhan and did not avoid traveling to Wuhan, even after the official confirmation of the person-to-person transmission. Finally, we should also point out that the coefficient estimates for *Treat* * *Before*_1,*t*_ are all statistically insignificant, which suggests that the parallel trend assumption for the DIDs are plausible.

#### Model 3: Wuhan 2020 vs. Seven Other Lockdown Cities2020

In Model 2, the coefficient estimates of *Treat*\**After*_*t*_ provide us with an estimate of the *sum* of the lockdown and the virus effects. In order to isolate the lockdown effect from the virus effect, we consider Model 3, where the estimation sample consists of data of the city of Wuhan and *seven other cities* that went into partial lockdown on February 2 and February 4, 2020, 12 to 14 days after the lockdown of Wuhan, in an effort to curtail the spread of the virus.^16^ As we show in Table A2 in the Appendix, these seven cities are more comparable to Wuhan than other unlocked cities in terms of the epidemic situation and other economic indicators, and thus provide a reasonable control group to partial out the virus effect. In particular, it is much more plausible than in Model 1 (where the control cities are cities that were never locked down) to assume that the deterrence effect of the virus on human mobility in the seven cities is similar to that in Wuhan.

The estimation sample for Model 3 consists of data from Wuhan and the seven cities for the period between January 1 and February 2, 2020. Note that during this period, none of the seven control cities were locked down yet, even though they were soon eventually locked down. The definition for *Treat* variables are as follows. In Panel A, *Treat* takes value 1 if the destination city *i* is Wuhan; in Panel B, *Treat* takes value 1 if the origination city *j* is Wuhan; in Panel C, *Treat* takes value 1 if city *i* is Wuhan. The control group consists of the seven cities.

Under Model 3, the coefficient estimate of *Treat* * *Before*_2,*t*_ measures the panic effect related to the virus outbreak in Wuhan *relative to* the seven control cities; and the estimate of *Treat*\**After*_*t*_ measures the *lockdown* effect only. The coefficient estimate of *Treat*\**Before*_1,*t*_ still allows us to examine whether the parallel trend assumption for DID is satisfied. The possibly time-varying Spring Festival effects and the virus effects are both absorbed in the day fixed effects.

We find that the Wuhan lockdown significantly reduces the inflow into, outflow from, and within-city movements in Wuhan by 76.64% (= 1 − *exp*(−1.454)), 56.35% (= 1 −*exp*(−0.829)), and 54.16% (= 1 − *exp*(−0.780)), respectively. We interpret these as the *pure* lockdown effect on population mobility related to Wuhan.

#### Summary

Based on our preferred estimation models, which are models 2 and 3, Table 3 summarizes our estimates of the panic effect, the virus effect, and the lockdown effect on inflows into, outflows from Wuhan, and within-city population movements in Wuhan.

**Table 3:** Summarizing the Panic Effect, Virus Effect and Lockdown Effect on Inter-City and Within-City Population Movements of Wuhan

| Effect | Infows | Outflows | Within-City |
| --- | --- | --- | --- |
| Panic Effect | -11.48% | +107.09%*** | -24.19%*** |
| Virus Effect | -65.63%*** | -36.75%*** | -66.41%*** |
| Lockdown Effect | -76.64%*** | -56.35%*** | -54.15%*** |

*Notes:* These effects are calculated based on the estimates reported in Columns (2) and (3) of Table 2. \*\*\* Significant at the 1 percent level. \*\* Significant at the 5 percent level. \* Significant at the 10 percent level.

In Table 3, the lockdown effects are directly calculated from the corresponding coefficient estimates of *Treat* * *After* from Model 3 discussed above; the panic effects are from the coefficient estimates of *Treat* * *Before*_2_ in Model 2. For the virus effect, we recognize that the coefficient estimates of *Treat* * *After* in Model 2 incorporate both the lockdown and the virus effects. Thus we calculate the *virus effect* on inflows into Wuhan to be *exp*(−2.522 − (−1.454)) − 1 = −65.63%, on the outflows from Wuhan to be *exp*(−1.287 − (−0.829)) − 1 =−36.75%, and on the within-city flow in Wuhan to be *exp*(−1.871−(−0.780))−1 = −54.15%.

Because our models assume that the different effects enter exponentially in explaining the flows — recall the natural log specifications in Equations (1) and (2— when we would like to calculate the impact of two or more effects on the population flows, we should not simply add the individual effects. For example, the joint impact of the panic and virus effects on outflows out of Wuhan is (1 + 107.09%) * (1 − 36.75%) − 1 = 30.98%, instead of the simple sum of 107.09% − 36.75% = 70.34%.

## 4 Quantify the Impact of Lockdown on the National Spread of COVID-19

### 4.1 Inter-city Flow and the 2019-nCoV Transmission

We now examine the impact of human mobility on the transmission of 2019-nCoV. Considering that almost all the new COVID-19 cases outside the city of Wuhan were confirmed after the Wuhan lockdown while almost all inter-city population flows occurred prior to the Wuhan lockdown (see Figures 2 and 3), we investigate the imported infections by specifically looking at the impact of population inflows from cities in the epicenter of the Novel Coronavirus outbreak, namely, Wuhan and other cities in Hubei province, on the new cases in the destination cities.

**Figure 2:**
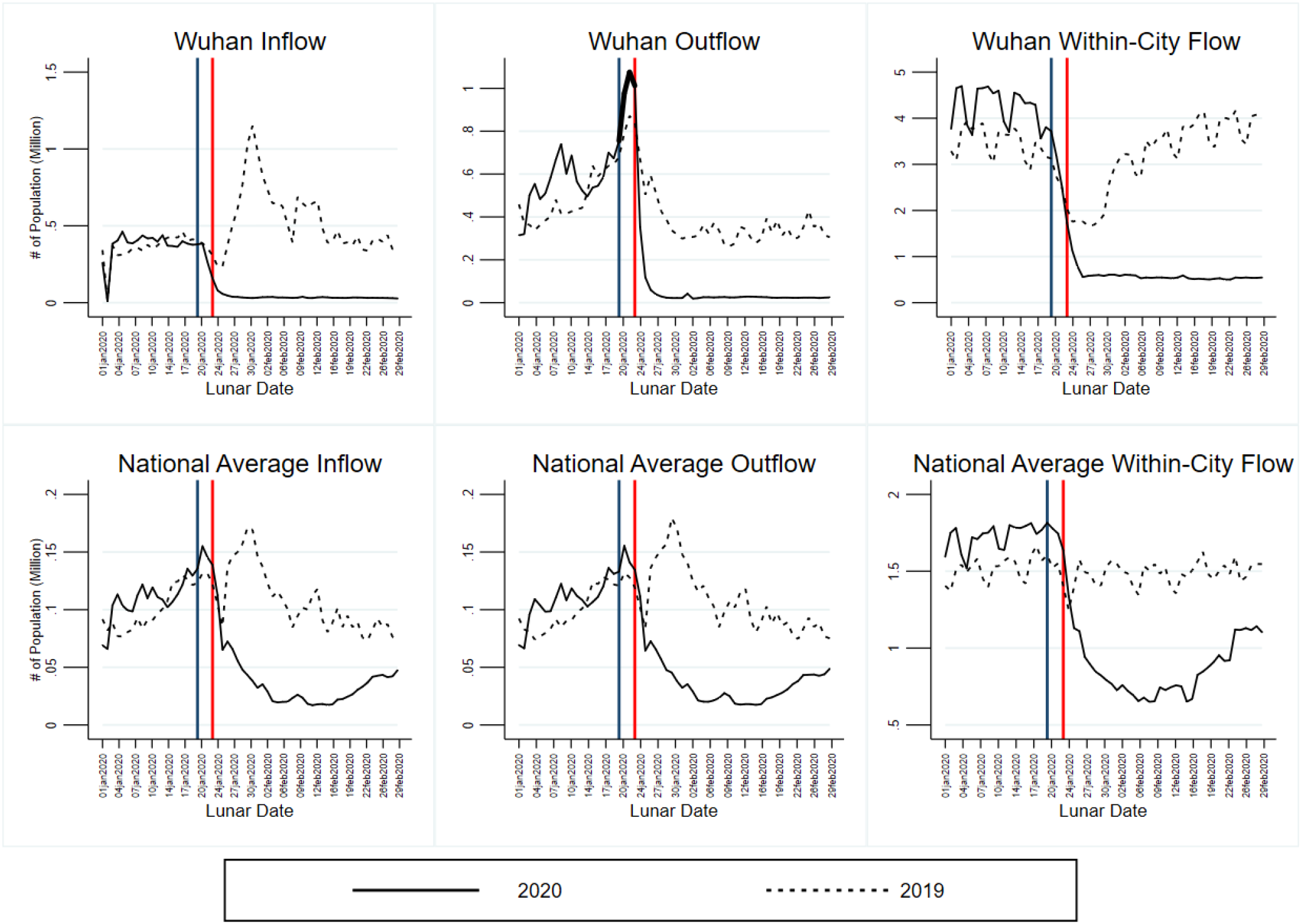
Inter-city and Within-city Population Flows. *Notes*: This figure presents the daily inflow into, outflow from, and within-city flow in Wuhan (top figures) and in other cities in China (bottom figures) during the sample lunar calendar date matched period in 2019 and 2020. The blue line indicates the date of official confirmation of human-to-human spread of COVID-19 (January 20, 2020) and the red line indicates the date of Wuhan lockdown (January 23, 2020).

**Figure 3:**
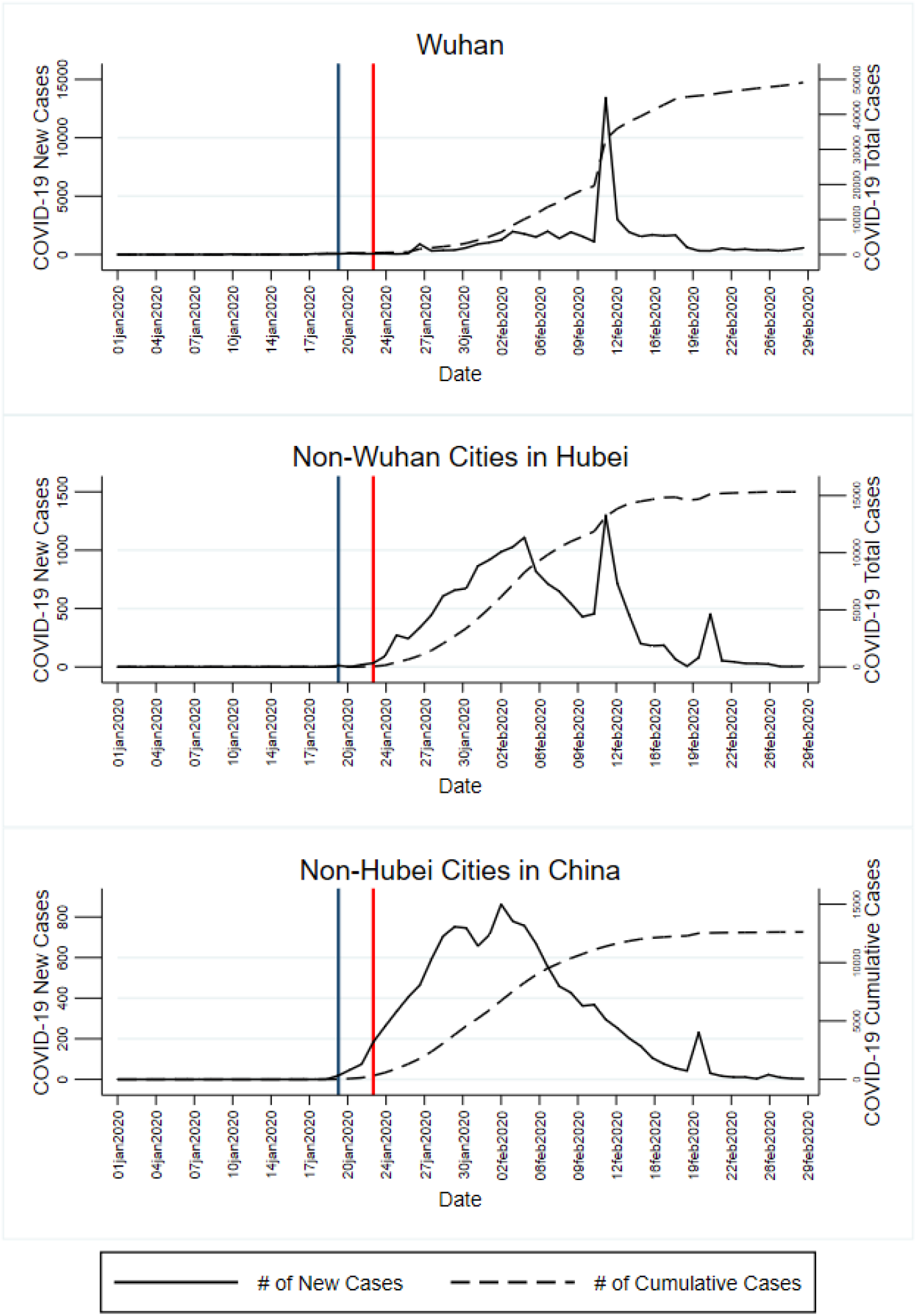
Daily Confirmed Cases and Cumulative Confirmed Cases. *Notes*: This figure shows the daily confirmed, dead, and healed cases in Wuhan, other cities in Hubei Province, and cities outside of Hubei Province.

Recognizing that 2019-nCoV has a long incubation period, we estimate a *dynamic distributed lag* regression model taking into account that inflows from Wuhan with different lags may have differential impacts on the current new cases in the destination cities. Most of the medical literature states that the 2019-nCoV virus has a median incubation period of five days, and some can have an incubation period of 14 days or more (see Lauer et al. (2020), e.g.). Luckily, our data allows us to incorporate the possibility that contact with an infected person from Wuhan or other cities in Hubei can result in confirmed infections in the destination city for up to 22 days.

The analysis focuses on the daily new confirmed COVID-19 cases in the post-Wuhan lockdown period from January 23 to February 29, 2020, for cities *i* that are *outside of Hubei province*. Specifically, we run the following regression:^17^

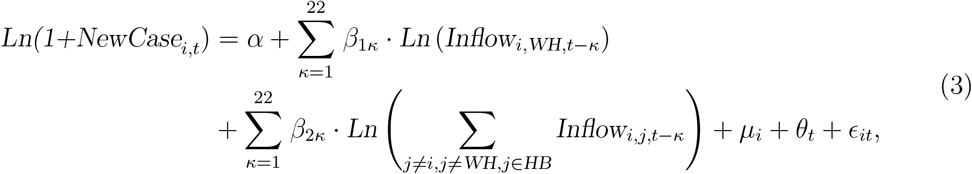

where *i* indexes the cities outside of Hubei, and *t* ∈ {23, …, 60} indicate the date.^18^ *κ* ∈{1, …, 22} indicates the time lapsed from the inflows from Wuhan or other Hubei cities till the current date *t. Ln(1+NewCase*_*i,t*_) is the logarithm of the number of new confirmed cases in city *i* at date *t. Inflow*_*i,WH,t*−*κ*_ and ∑_*j* ≠*i,j* ≠*WH,j*∈*HB*_ *Inflow*_*i,j,t*−*κ*_ are the inflows from Wuhan, and the inflows from the 16 other cities in Hubei to city *i*, respectively, *κ* days prior to the focal date *t*. We control for destination city fixed effects *µ*_*i*_ and date fixed effects *θ*_*t*_.

Note that, in this regression we include only cities outside of Hubei Province for two reasons. First, Wuhan and other cities in Hubei province are the epicenter of the Novel Coronavirus outbreak, and we are interested in how population outflows from these cities to other cities outside Hubei affect the destination cities’ COVID-19 cases. Second, the confirmed COVID-19 cases in Wuhan and other cities in Hubei province are likely to be inaccurate for the following reasons. First, as widely reported, the health care systems in Wuhan and other cities in Hubei were totally overwhelmed by the sheer number of COVID-19 patients. This made it impossible to conduct laboratory tests on all patients, which can lead to delayed confirmation of infected patients. Second, during the delay some of the infected may have healed on their own, or have died before being confirmed. Third, local government officials may face strong incentives to under-report the number of infected cases. In Section 4.2, we will evaluate the possible downward biases of the officially reported cases in Wuhan and other cities in Hubei based on our estimates. In contrast, the confirmed cases in other cities are likely to be accurate, as their numbers are not large enough to overwhelm their local health care system; and the incentives to under-report are much weaker in cities outside of Hubei.

The estimated coefficients *β*_1*κ*_ and *β*_2*κ*_ in Equation (3) respectively represent the impact of the inflows from Wuhan and other cities in Hubei *κ* ∈ {1, …, 22} days ago on the destination cities’ new cases today. They are respectively plotted in the top and bottom panels of Figure 4. We also fit a spline smoothed curve of the estimated effects of the different lags of inflows from Wuhan and Hubei, which both show a clear inverted *U* -shape with respect to the lags. Interestingly, both graphs show that the largest impact on the newly confirm cases today in Chinese cities outside Hubei comes from the inflow population from Wuhan or other cities in Hubei about 12 to 14 days ago. The pattern exhibited in Figure 4 is consistent with the hypothesis that the incubation period of the 2019-nCoV is up to 12 to 14 days, but also consistent with a shorter incubation period coupled with secondary infections.

**Figure 4:**
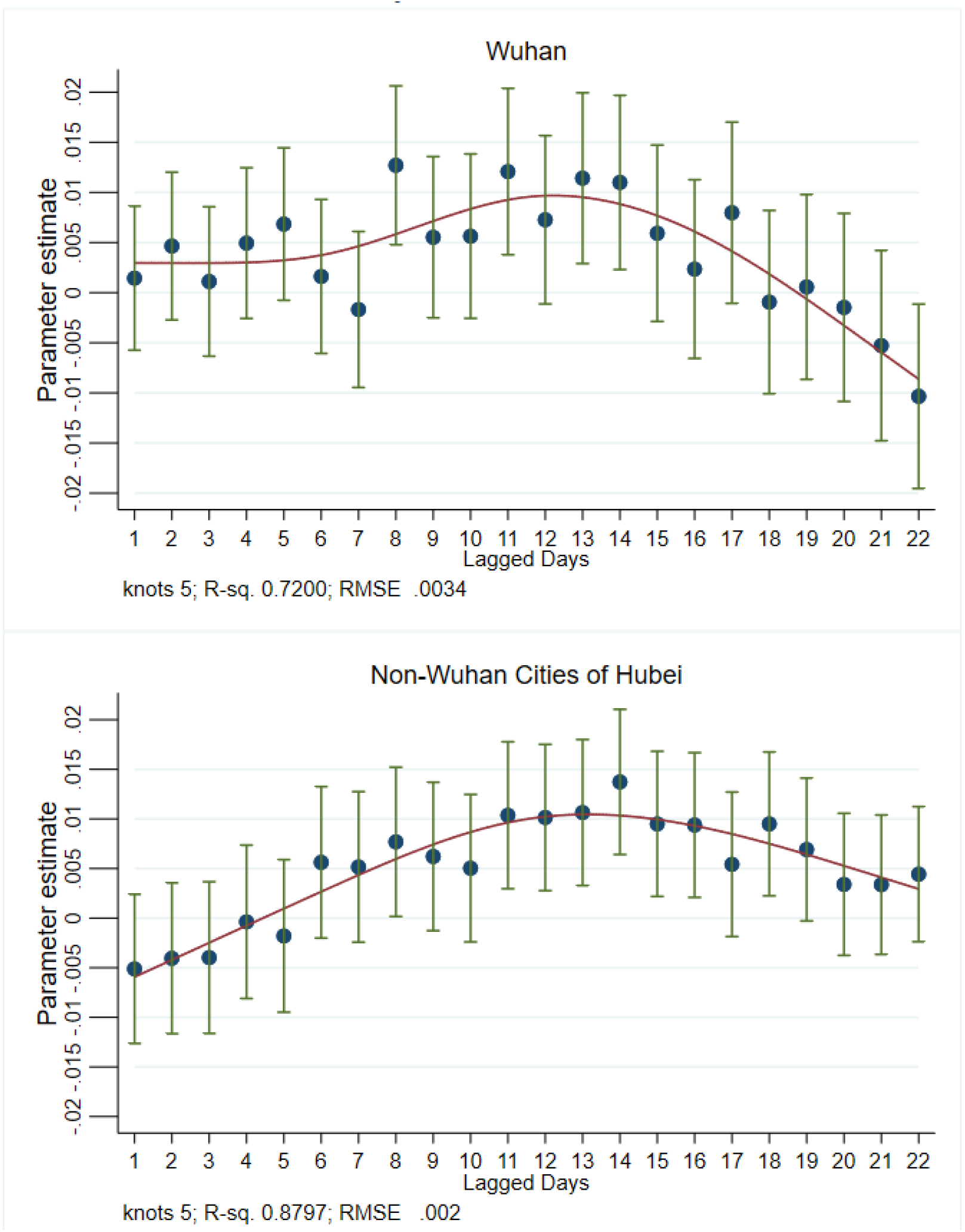
Dynamic Impact of Past Inflow from Wuhan and from Other Cities in Hubei on Daily New Cases. *Notes*: This figure plots the dynamic effects of lagged inflows from Wuhan (left) and 16 other cities in Hubei (right) from estimating Equation (3). We add spline smoothing fit curves (in red) using the *rcspline* function and plot the 95% confidence intervals (the vertical green whiskers).

### 4.2 Estimating the “Actual” Number of Infection Cases in Wuhan and Other Cities in Hubei

Anecdotal evidence suggests the official statistics of COVID-19 cases in Wuhan may have been under-reported due to the shortages of testing equipment and other medical resources. With the estimated dynamic effects as shown in Figure 4, which is estimated under the plausible assumption that the reported cases outside Hubei Province are reliable, we can estimate the “actual”‘ number of infection cases in Wuhan and other cities in Hubei.

To estimate the “actual” number of infection cases in Wuhan using the estimated Equation (3), we technically need to impute a value for *Inflow*_*WH,WH,t*−*κ*_, that is “inflows from Wuhan to Wuhan.” We proxy these inflows by the daily *within-Wuhan* population movement from January 1 to February 29, i.e., by *WithinCityFlow*_*WH,t*−*κ*_. Similarly, to estimate the “actual” number of infection cases in other cities in Hubei, we need to replace the inflow from city *j* to itself by the corresponding daily within-city-*j* population movements.

We need to make an additional assumption about the city fixed effects. Recall that cities in Hubei province were not included in the estimation sample for Equation (3), as such there are no city fixed effects estimated for cities in Hubei. Luckily, it is plausible to assume that the city fixed effects in Hubei is the average of the city fixed effects of all Chinese cities outside Hubei. According to World Bank’s (respectively, IMF’s) method of estimating per capita GDP, Hubei’s per capita GDP in 2018 was USD 10,067 (respectively, USD 10,054), and the average per capita GDP of mainland China was USD 9,769 (respectively, USD 9,750). Hubei is not only geographically and demographically in the center of mainland China, it is also economically the average of China. Thus we believe it is defensible to proxy the fixed effects of Hubei cities by the average of the city fixed effects outside of Hubei.

In Figure 5, we plot the estimated daily new cases according to the above-described method using the estimated Equation (3), as well as the corresponding cumulative cases for Wuhan (Panel A) and 16 other cities of Hubei (Panel B) for the period of January 23 to February 29, 2020. We also plotted the corresponding daily and cumulative officially reported (i.e., documented) cases.

**Figure 5:**
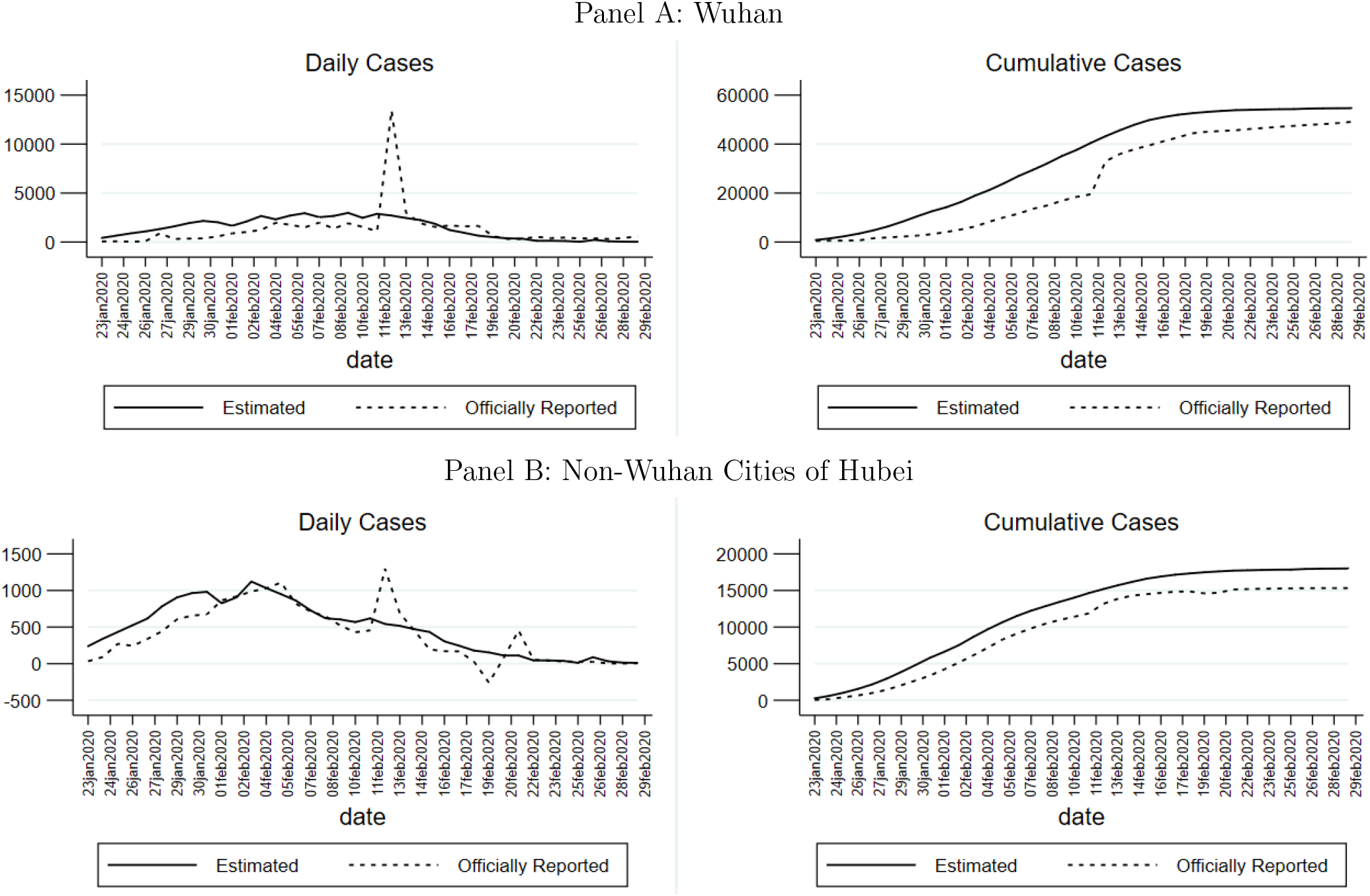
Estimation on the “Actual” Number of Infected Cases in Wuhan and No-Wuhan Cities in Hubei. *Notes*: This figure compares the estimated COVID-19 cases (in solid curve) with the officially reported confirmed cases (in dotted curve) in Wuhan (top) and in 16 non-Wuhan cities in Hubei Province (bottom) from January 23 to February 29 in 2020. The left panel plots the estimated new COVID cases on each date *t* from 23 (January 23, 2020) to 60 (February 29, 2020) obtained from Equation (3). The right panel plots the estimated cumulative cases on each day.

We find a persistent gap between the estimated and reported laboratory-confirmed cases in Wuhan before February 11, 2020, just before the announcement of a new Party Secretary for Hubei province on February 12, 2020. The estimated “actual” number of infection cases is 2.48 times the reported cases during the first 20 days after the Wuhan lockdown, on average. In particularly, we estimate that on January 23, 2020, the day of the Wuhan lockdown, 42.17% of our estimated infections in Wuhan were undocumented in the sense that the number of officially reported cases on that day was only 57.83% of our estimated infection cases. This gap widened over time, possibly as a result of the overwhelmed health care system, and peaked at 80.03% on January 26. The proportion of undocumented infections started to decline gradually afterward, when more medical support and resources were mobilized across China to support Wuhan. As of February 29, we estimate that there were 54,689 total COVID-19 infections in Wuhan, which is 11.33% higher than the official reported statistics for Wuhan — a total of 49,122 cases. The 11.33% discrepancy can be plausibly be explained by the unaccounted for self-healing and death that might have occurred during the early periods of the outbreak between January 23 and early February. Thus, we are led to conclude that the almost all infection cases in Wuhan were able to be treated over time as the stress on the health system was relieved, and moreover, the official statistics were mostly accurate, as can be seen from the left figure on the daily new cases in Panel A in Figure 5.

It is useful to note that the general pattern of the undocumented COVID-19 infections in Wuhan before and after the lockdown based on our estimates is consistent with that reported in Li et al. (2020). They use a networked dynamic meta-population model and Bayesian inference to estimate the proportion of undocumented infections in the epicenter of the outbreak, as well as the respective inflection rates for documented and undocumented cases. They estimate that 86% of all infection cases as of January 23, 2020 were undocumented. Our interpretation of these undocumented cases is broader than the asymptomatic cases (i.e. COVID-19 cases that do not show symptoms); we believe that some of the undocumented cases were due to the lack of ability to test the rapidly increasing infection cases in the early period of the outbreak. Most of the undocumented cases become documented over time as the care capacity was strengthened in Wuhan.

In the bottom panel of Figure 5, we plot our estimated daily new confirmed cases and total infection case for 16 cities (other than Wuhan) in Hubei province, together with the officially reported series. We find that in the 16 cities, infections were more seriously under-reported in the first week after the Wuhan lockdown when our estimated infected cases are 1.87 times of the reported cases. Our estimate reveals a very high rate of undocumented infections on the first day of Wuhan lockdown: 79.8%. The gap narrowed gradually with more medical resources provided and more stringent control measures implemented in those cities. By the end of our study period on February 29, 2020, the estimated “actual” number of infections is 17,999 cases in 16 other cities in Hubei, which is 17.4% higher than the officially reported cumulative cases (15,330). Again, the discrepancy between the estimated and officially reported cumulative cases can at least be partially attributed to the unaccounted for self-healing and death that might have occurred during the early periods of the outbreak.

### 4.3 How Many COVID-19 Cases Were Prevented by the Wuhan Lockdown?

Locking down Wuhan, a city of 11 million residents, was an unprecedented measure to contain the spread of the Novel Coronavirus. An important policy-relevant question is, then, how many COVID-19 cases were actually prevented by the Wuhan lockdown in China? To answer this question, we must estimate the *counterfactual* number of COVID-19 cases that would have occurred in other cities in the absence of Wuhan lockdown, which would, in turn, require a counterfactual estimate of the outflows from Wuhan to other Chinese cities, had there been no lockdown of Wuhan. In Section 3, we provided the estimates of how panic effect and virus effect will separately affect the outflows from Wuhan to other cities in China, separate from the lockdown effect. These effects are summarized in Table 3. It suggests that, in the absence of the Wuhan lockdown, the virus effect and panic effects would have led to a 36.75% decrease and a 107.09% increase in the outflow population from Wuhan, respectively. Based on these effects, Therefore, in the absence of Wuhan lockdown, we would expect that the outflows from Wuhan in days after January 23 – the date of Wuhan lockdown – to be

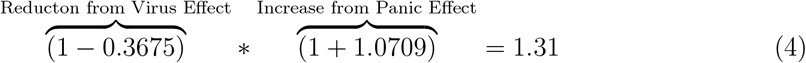

times higher than the *normal outflows* from Wuhan to other cities.

We use the daily level of outflows from Wuhan to a city *in 2019 on the same lunar calendar day* as a measure of the normal outflow, and multiple the number by 1.31 to obtain the daily counterfactual inflows from Wuhan to the city, had there been no lockdown of Wuhan from January 23, 2020. Using this estimation method, we find that on average, the estimated counterfactual outflows from Wuhan to 16 other cities in Hubei between January 23 and February 29, 2020 would be 16,568,984, a level that is 5.97 times the actually observed inflow population observed in the data in 2020 (with the lockdown), which is 2,774,793; similarly, the average inflows from Wuhan to the 347 other cities outside Hubei in China would be 2,274,455, 3.14 times the actual inflow population to those cities during the same period in 2020, which is 723,574.^19^

We denote the counterfactual inflows from *j* = *Wuhan* into city *i* at date *s* ∈ {23, …, 60} from the above calculation as 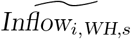 We assume that the Wuhan lockdown did not impact the population movements within other cities, and population flows among other cities. We also assume that all the control measures implemented by other cities after the Wuhan lockdown remain in place. Thus the parameter estimates of the dynamic lag effects of inflows from Wuhan and other cities in Hubei, estimated in Equation (3), remain valid as an epidemiological diffusion equation that is not affected by human mobility restrictions that result from the Wuhan lockdown; the lockdown only affected the human flows.

With these considerations in mind, we simulate the counterfactual number of COVID-19 cases, had there been no Wuhan lockdown, on date *t* ∈ {23, …, 60} (i.e., from January 23 to February 29, 2020) in cities *i* outside Hubei province by the following equation:

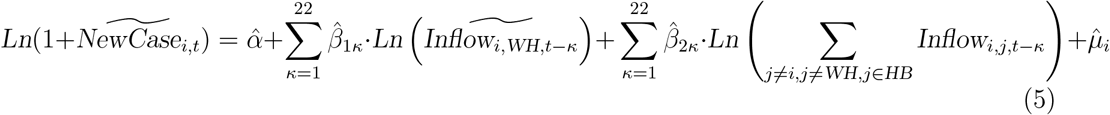

where 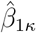 and 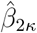 are coefficient estimates obtained from regressions specified in Equation n(3) and reported in Figure 4, and and 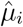 are the estimated city fixed effects from the same regression. Note that in predicting the counterfactual COVID-19 cases without Wuhan lockdown, we use the *counterfactual* inflows from Wuhan to city *i* for days after January 23 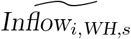 discussed previously.

We are also interested in predicting the counterfactual cases in other non-Wuhan cities in Hubei province using the same method as that described above, with two differences. First, since Hubei cities are not included in the estimation of Equation (3), we do not have the city fixed effects for the Hubei cities. As we argued in Section 4.2, it is plausible to assume that the fixed effects of Hubei cities are the average of city fixed effects of the cities outside Hubei. We maintain this assumption as well in this counterfactual exercise. Second, following the same strategy of Section 4.2, for a non-Wuhan city *j* in Hubei province, we use the within-city-*j* population movement at date *t* to proxy for the inflow to city *j* from city *j* when we implement Equation (5) for non-Wuhan Hubei cities.

In Figure 6, we present the counterfactual estimates of COVID-19 cases had there been no Wuhan lockdown (in solid curve), and the officially reported cases (in dashed curve) for cities outside Hubei province (Panel A) and non-Wuhan cities of Hubei (Panel B). The left figure in each panel depicts the model’s counterfactual prediction and the actual of daily infection cases, and the right figure depicts the evolution of cumulative cases from January 23 to February 29, 2020.

**Figure 6:**
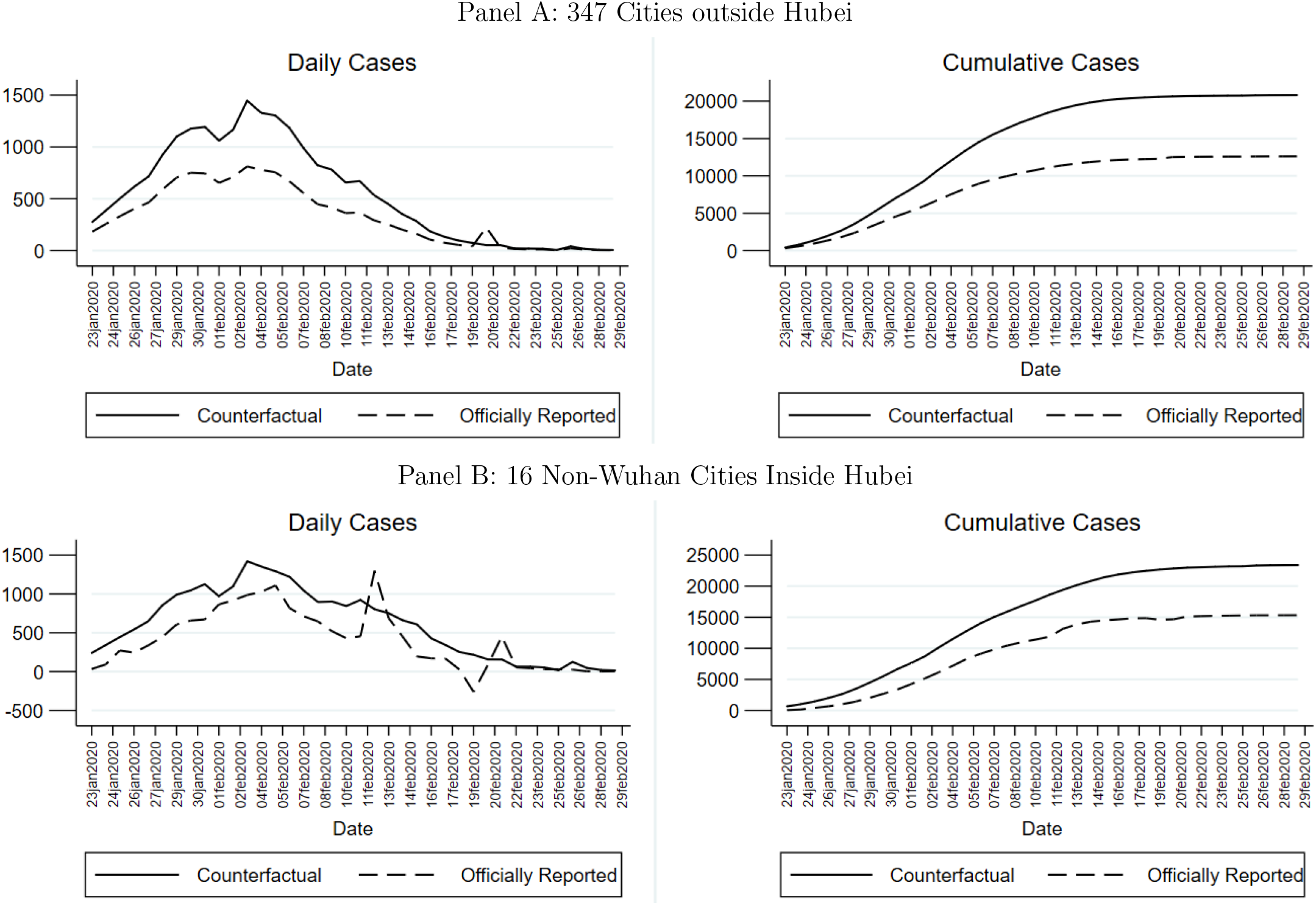
Counterfactual of Infected Cases Elsewhere in China If Wuhan Were Not Locked Down from January 23, 2020. *Notes*: This figure plots the counterfactual estimation on the COVID-19 cases in 16 other cities outside Hubei (top) and 347 other cities in China (bottom) if Wuhan had never been under a government-ordered lockdown (in solid curve), and traces the officially reported COVID-19 cases in cities outside of Hubei (in dotted curve). The left graphs present the daily new cases and the right graphs present the cumulative daily cases.

The gap between the estimated counterfactual number of infection cases (the solid curve) and the officially reported cases (the dashed curve) on the right figure represent the number of COVID-19 cases prevented by the Wuhan lockdown. As of February 29, 2020, the officially reported number of COVID-19 in the 347 cities outside Hubei province was **12**,**626**, but our counterfactual simulation suggests that there would have been **20**,**810** cases, had there been no Wuhan lockdown. Similarly, the officially reported number of COVID-19 cases in the 16 non-Wuhan cities in Hubei was **15**,**330** by February 29, 2020, but our counterfactual simulation suggests that the number of infection cases would have been **23**,**400**, had there been no Wuhan lockdown. That is, the COVID-19 cases would be **64**.**81%** higher in 347 cities outside Hubei, and **52**.**64%** higher in 16 other cities in Hubei as of February 29, in the counterfactual world in which the city of Wuhan were not locked down from January 23, 2020. Our findings thus suggest that the lockdown of the city of Wuhan from January 23, 2020 played a crucial role in reducing the imported infections in other Chinese cities and halts the spread of 2019-nCoV virus.

Our model also allows us to project when the new infection cases would have peaked in other cities in the absence of Wuhan lockdown. As shown by the solid curves in the left figures of each panel, our model projects that in the absence of Wuhan lockdown, the new daily infection cases in the 16 non-Wuhan cities in Hubei would have peaked on February 2, the 9th day after the lockdown of Wuhan, at a level of 1,466 daily new cases; and elsewhere in China, it would have peaked on February 3, 2020, the 10th day after the lockdown of Wuhan, at a level of 1,082 daily new cases. We also find that the estimated daily new cases in the counterfactual world gradually converge to the reported daily cases from February 22, suggesting that the social distancing measures implemented elsewhere in China would have worked eventually to contain the spread of 2019-nCoV virus, even if the city of Wuhan was not locked down on January 23 2020, but the initial onslaught on the medical system in all cities in China would have been much more severe, and the total number of infection cases elsewhere would have been significantly higher.

## 5 Effect of Social Distancing on Virus Transmission in Destination Cities

As the 2019-nCoV virus spread throughout the world, many countries are also implementing lockdown measures, and mandate *social distancing* as a policy response to contain the spread of the virus. Up to now, our analysis has focused on the events of the lockdown of Wuhan – the epicenter of the Novel Coronavirus outbreak, it would also be interesting to study the impact of lockdowns and/or the social distancing measures in the destination cities in reducing and containing the spread of virus. Indeed, Chinazzi et al. (2020) point out that travel restrictions to and from Mainland China impact the global pandemic of the COVID-19 only if transmission within the community is simultaneously reduced by 50% or more, which suggests that social distancing at the destination cities is crucial in preventing the possibly asymptomatic transmission from the source city. Moreover, quantifying the effect of social distancing on virus transmission is especially relevant to the stage of pre-epidemic community spread when a person who is not known to have traveled to affected countries, or to have had contact with an infected person becomes infected.

As shown in Table A1 in the Appendix, within the few weeks after the Wuhan and Hubei lockdowns, various human mobility restrictions were imposed on 63 other Chinese cities outside Hubei. As described in Table A1, the “lockdowns” in destination cities varied in their degree of strictness, from building entrances checkpoints to establishing quarantine zones, and from public transit shutdowns to strict limits on the inflows into the city, outflows out of the city, as well as within-city population movements. We interpret the human mobility restrictions in the destination cities as an *enhanced social distancing* policy, because the “lockdown” rules in the destination cities are not as strict as those implemented in Wuhan. In this section, we use the variations in the destination cities’ “lockdown” policies to study how the effects of inflows from Wuhan and other cities in Hubei province on the spread of COVID-19 cases in the destination cities are impacted by the changes in the destination cities’ lockdown policies, which would drastically impact the within-city population movements in the destination cities.^20^ Specifically, we estimate the following specification that is a modified version of the regression specification described by Equation (3):

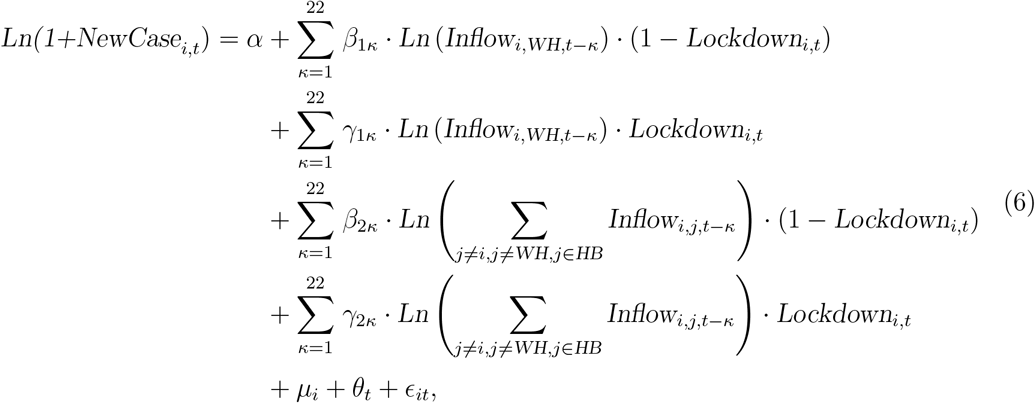

where the new variable *Lockdown*_*i,t*_ is a dummy that takes value 1 if time *t* is a date after destination city *i*’s “lockdown” date, if at all; and 0 otherwise, where the lockdown dates of the 63 cities outside Hubei are listed in Table A1. If city *i* never implemented any formal lockdown policy, the dummy is always 0. Therefore, the coefficients, *β*_1*κ*_ and *β*_2*κ*_, respectively, measure the impact of the lagged inflows from Wuhan and Hubei *κ* days earlier on the destination cities’ current new cases before the city’s imposition of its “lockdown,” while *γ*_1*κ*_ and *γ*_2*κ*_ represent the effect on destination cities’ of Wuhan and Hubei inflows after the imposition of the city’s lockdown. If enhanced social distancing that comes from the “lockdown policies” imposed at the 63 Chinese cities outside Hubei is effective in reducing the spread of the virus from population flows from the epicenter of the virus, then we expect that *γ*_1*κ*_ and *γ*_2*κ*_ to be smaller than *β*_1*κ*_ and *β*_2*κ*_, respectively.

In Figures 7 (respectively, Figure 8), we plot the estimated coefficients, *β*_1*κ*_ and *γ*_1*κ*_ (respectively, *β*_2*κ*_ and *γ*_2*κ*_) in Panel (a), and their differences in Panel (b) for the lagged effects of inflows from Wuhan (respectively, 16 non-Wuhan cities in Hubei) on the daily new cases in destination cities outside Hubei province. We find that the estimated lagged effects of inflows from Wuhan and other cities in Hubei before the destination city’s lockdown, if any, show little change compared to the coefficients in Figure 4 however, the coefficient estimates of lagged inflows after the destination city’s lockdown policies appear to be insignificant and indifferent from zero on almost all lags. In Panel (b) of Figures 7-8, we plot the differences between the estimated effects pre and post the destination cities’ lockdown policies. We find that the differences between the estimated coefficients pre and post destination city lockdowns are positive and statistically significantly at at-least 10% level for 7 (respectively, 5) of the first ten lagged population inflows from Wuhan (respectively, 16 other cities of Hubei), and the other differences in lagged estimates are statistically insignificant.

**Figure 7:**
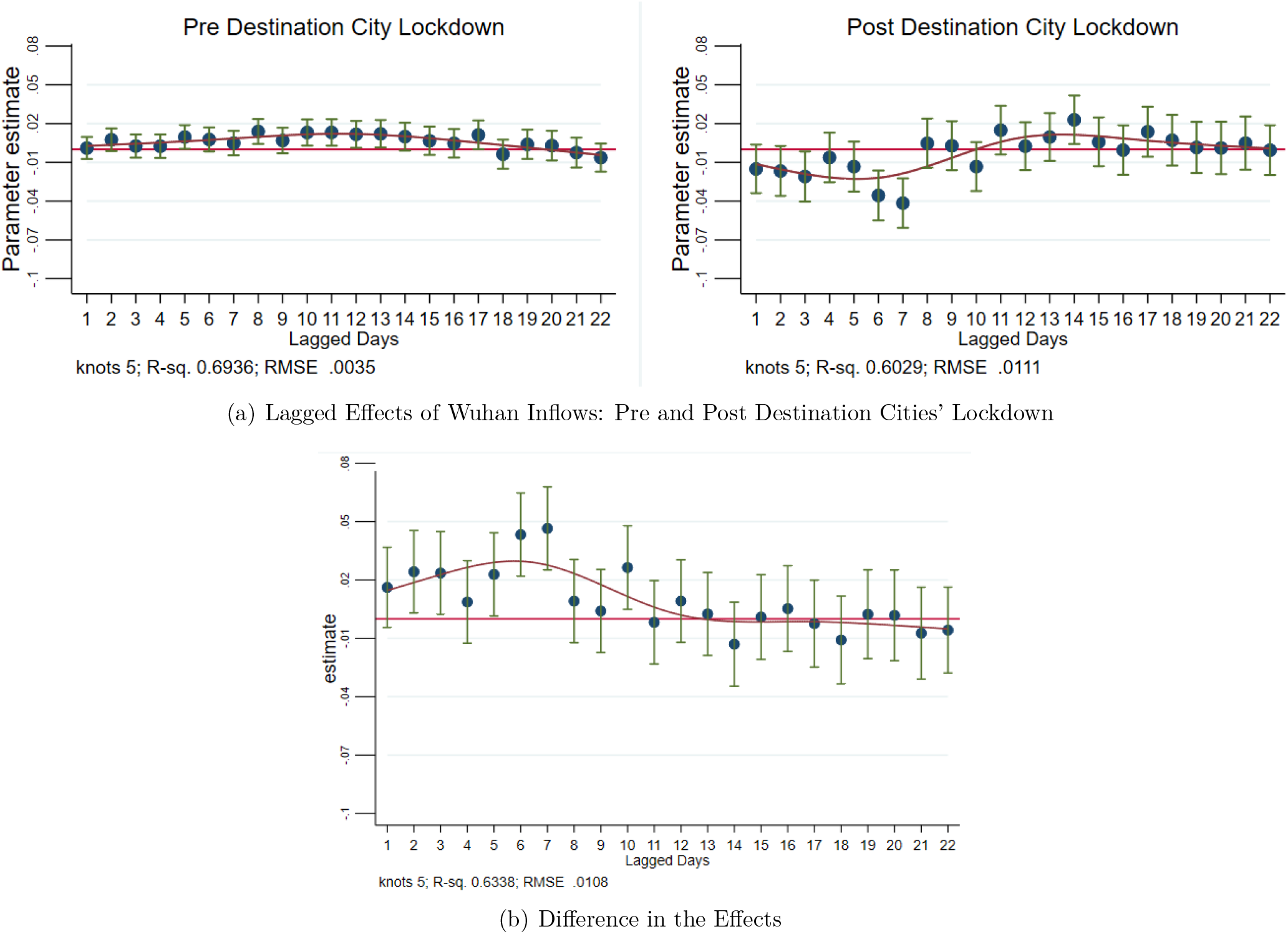
Dynamic Impact of Past Inflows from Wuhan on Destination Cities’ Daily New Cases: Pre and Post Destination Cities’ Lockdown. *Notes*: This figure plots the dynamic lagged effects of past inflows from **Wuhan** pre (left figure of Panel (a)) and post (right figure of Panel (a)) destination cities’ lockdown policy, if any. Panel (b) plots the difference of the pre- and post-estimated effects. The coefficient estimates are obtained from estimating Equation (6). We add spline smoothing fit curves using the *rcspline* function and plot the 95% confidence intervals (the vertical green whiskers).

**Figure 8:**
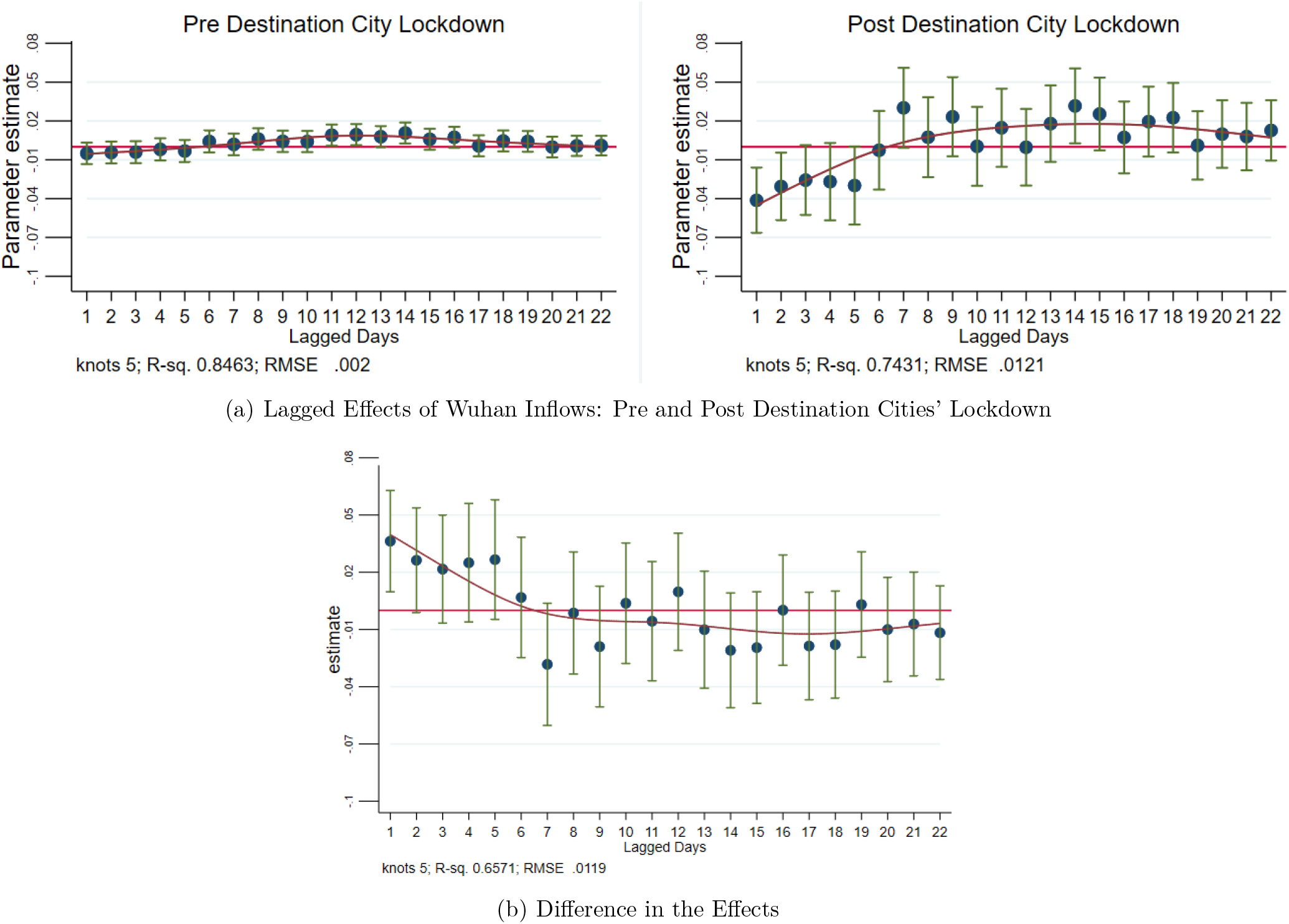
Dynamic Impact of Past Inflows from Non-Wuhan Hubei Cities on Destination Cities’ Daily New Cases: Pre and Post Destination Cities’ Lockdown. *Notes*: This figure plots the dynamic lagged effects of past inflows from 16 **non-Wuhan Hubei cities** pre (left figure of Panel (a)) and post (right figure of Panel (a)) destination cities’ lockdown policy, if any. Panel (b) plots the difference of the pre- and post-estimated effects. The coefficient estimates are obtained from estimating Equation (6). We add spline smoothing fit curves using the *rcspline* function and plot the 95% confidence intervals (the vertical green whiskers).

These results suggest that the enhanced social distancing policies in the destination cities are effective in reducing the impact of population inflows from the source cities of Wuhan and other cities in Hubei on the spread of 2019-nCoV virus in the destination cities. This in turn implies that population inflows from the epicenter contribute to the spread of infection in the destination cities only *before* the social distancing measures are applied; it appears that after implementing their various control measures, cities adopting an extended lockdown can flatten the upward trajectory of the virus.

## 6 Conclusion

In this paper, we quantify the causal impact of human mobility restrictions, particularly the lockdown of the city of Wuhan on January 23, 2020, on the containment and delay of the spread of the Novel Coronavirus, and estimate the dynamic effects of up to 22 lagged population inflows from Wuhan and other Hubei cities, the epicenter of the 2019-nCoV outbreak, on the destination cities’ new infection cases. We find that the lockdown of Wuhan reduced inflow into Wuhan by 76.64%, outflows from Wuhan by 56.35%, and within-Wuhan movements by 54.15%. Using counterfactual simulations with these estimates, we find that the lockdown of the city of Wuhan on January 23, 2020 contributed significantly to reducing the total infection cases outside of Wuhan, even with the social distancing measures later imposed by other cities. We find that the COVID-19 cases would be 64.81% higher in the 347 Chinese cities outside Hubei province, and 52.64% higher in 16 non-Wuhan cities inside Hubei, in the counterfactual world in which the city of Wuhan were not locked down from January 23, 2020. We also find that there were substantial undocumented infection cases in the early days of the 2019-nCoV outbreak in Wuhan and other cities of Hubei province, but over time, the gap between the officially reported cases and our estimated “actual” cases narrows significantly. We also find evidence that imposing enhanced social distancing policies in the 63 Chinese cities outside Hubei province is effective in reducing the impact of population inflows from the epicenter cities in Hubei province on the spread of 2019-nCoV virus in the destination cities elsewhere.

The results from our analysis provide valuable causal evidence on the role of human mobility restrictions on the containment and delay of the spread of contagious viruses, including the 2019-nCoV virus that is now ravaging the world. Enhanced social distancing in the destination cities, and, if an epicenter can be identified as was the case for the city of Wuhan in China, a lockdown, can play crucial roles in “flattening” the daily infection cases curve, giving the stressed medical system a chance to regroup and deal with the onslaught of new infection cases. Although our study focuses exclusively on the effect of human mobility restrictions on the spread of 2019-nCoV virus in China, our estimated results can have general implications to other countries in their fight against the Novel Coronavirus.

## Data Availability

We obtain inter-city population migration data from Baidu Migration, a travel map offered by the largest Chinese search engine, Baidu (http://qianxi.baidu.com/);
COVID-19 daily case counts are collected from China CDC, which provides daily updates on confirmed, dead, and recovered COVID-19 cases in each cityFrom January 11 to February 29, 2020 (http://2019nCoV.chinacdc.cn/2019-nCoV/)

## Appendices

**Table A1.**
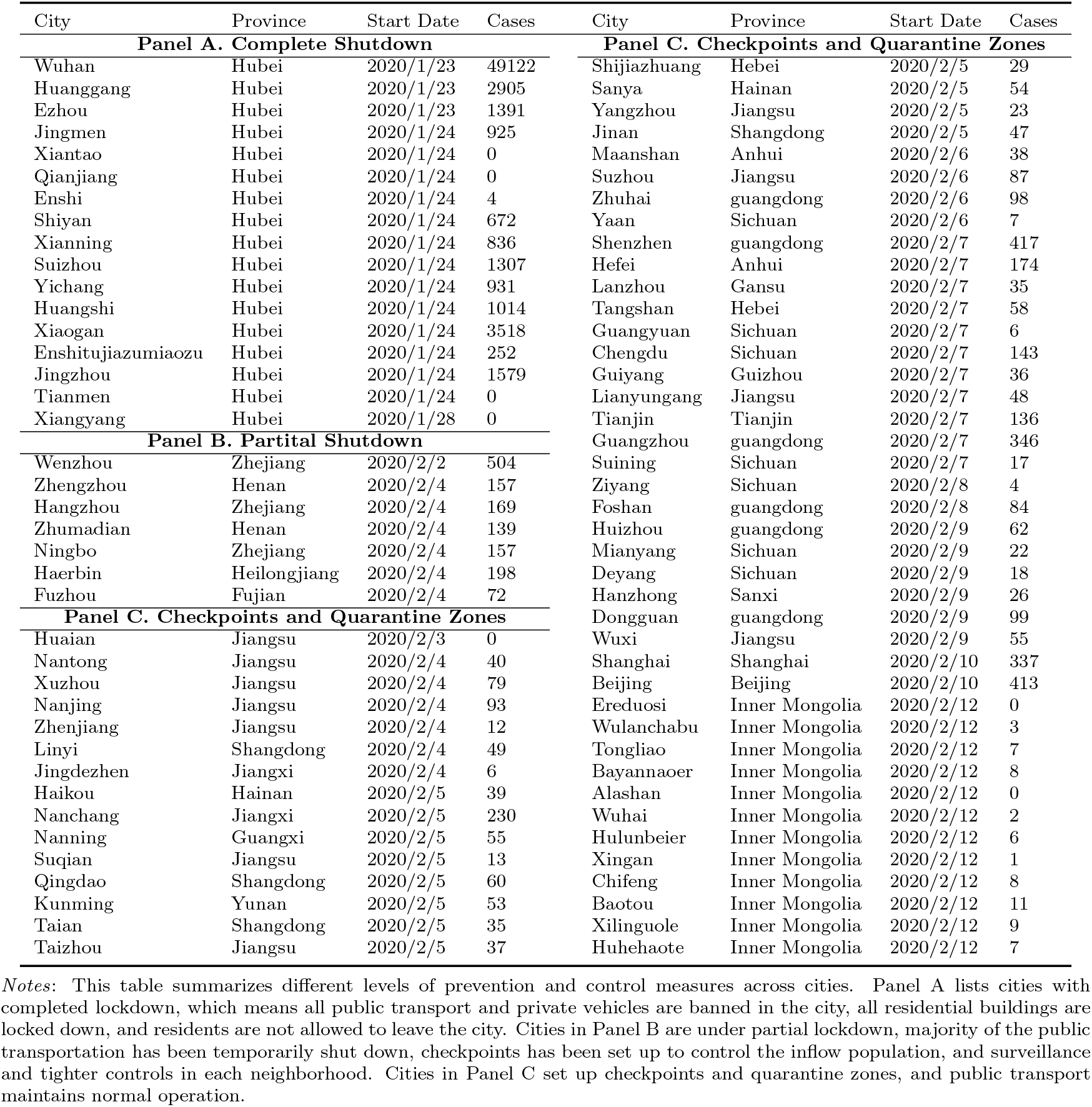
Various Levels of Prevention and Control Measures in Different Cities

**Table A2.**
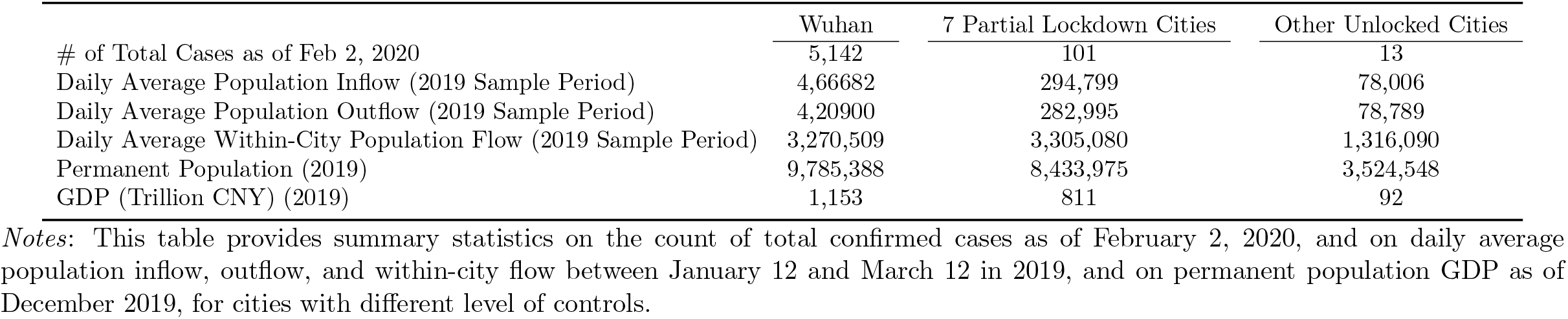
Summary Statistics of Cities with Different Level of Controls

## Footnotes

1 Throughout the paper, we use 2019-nCoV as the official name for the Novel Coronavirus according to the World Health Organization, and use COVID-19 as the name of the disease caused by 2019-nCoV.

2 Source: https://www.who.int/emergencies/diseases/novel-coronavirus-2019

3 With a population of over 11 million, Wuhan is the largest city in Hubei, the most populous city in Central China, and the seventh most populous city in China. The city is also a major transportation hub, with dozens of railways, roads and expressways passing through and connecting to other major cities, and home to 82 colleges and more than 1 million college students.

4 Baidu Migration uses Baidu Maps Location Based Service (LBS) open platform and Baidu Tianyan to calculate and analyze the LBS data, and provides visual presentation to show the trajectory and characteristics of the population migration. Source: http://qianxi.baidu.com/

5 Baidu has been the dominant search engine in China because all Google search sites have been banned in mainland China since 2010.

6 It is important to emphasize here that the mobility data is about the movement of people from one city to another based on geo-location services of the smartphones; as such a person flowing out of city A to city B is not necessarily a *resident* of city A, but he/she must have been to city A before moving to city B.

7 Source: http://2019nCoV.chinacdc.cn/2019-nCoV/

8 The spike of confirmed cases observed on February 12 in Hubei Province is. for the most part, the result of a change in diagnosis classification for which 13,332 clinically (rather than laboratory) confirmed cases were all reported as new cases on February 12, 2020, even though they might have been clinically diagnosed in the preceding weeks. Also on February 12, 2020, a new Communist Party Secretary of Hubei province was appointed who started his position on the next day.

9 The range of the incubation period for 2019-nCoV is estimated to be 2-14 days, or even as long as 24 days. The median incubation period is about 5 days, see Lauer et al. (2020). In contrast, the incubation period for SARS is 2-7 days. In addition, SARS transmits only after showing symptoms.

10 Within hours of the Wuhan lockdown, travel restrictions were imposed on the nearby cities of Huanggang and Ezhou, and were eventually imposed on all 14 other cities in Hubei, affecting a total of about 57 million people. On February 2, the city of Wenzhou implemented a partial lockdown in which only one person per household was allowed to exit once every two days, and most of the highway exits were closed. Following Wenzhou, another six cities, Hangzhou, Zhumadian, Ningbo, Haerbin, Fuzhou, and Zhengzhou also launched similar partial lockdowns on February 4. In another 56 cities, surveillance and tighter controls applied to each neighborhood. In the provinces of Liaoning and Jiangxi, as well as major cities such as Shenzhen, Guangzhou, Nanjing, Ningbo, Chengdu and Suzhou, checkpoints have been set up to control the inflow population.

11 The migration across China, which officially begins from about two weeks before, and ends about three weeks after, the Lunar New Year is often referred to as *Chunyun* (meaning Spring movement). In 2019, approximately 3 billions trips were made during Chunyun, see https://www.cnn.com/travel/article/lunar-new-year-travel-rush-2019/index.html.

12 The large outflow from Wuhan is possible because many people in Wuhan are migrant workers and college students with hometowns elsewhere.

13 On January 20, 2020, an expert at China CDC confirmed that the Novel Coronavirus can spread from human to human. The confirmation highlighted the increasing risk of an epidemic. Prior to the January 20 announcement, the experts’ assessment of the virus was that it was “preventable and controllable,” and “at this time, there is no evidence of person to person transmission.”

14 Recall that Baidu Migration data covers 364 Chinese cities, 80 cities implemented some level of mobility restrictions (see Table A1).

15 If the virus effect is stronger in Wuhan than in the unlocked control cities, then the coefficient estimate of *Trea* \**After*_*t*_ also includes the excess virus effect on the population in Wuhan over the average virus effect experienced by the population in the control cities.

16 These seven cities are: Wenzhou, which was partially lockdown from February 2, 2020; and Ningbo, Zhumadian, Hangzhou, Zhengzhou, Haerbin, and Fuzhou, which were partially locked down on February 4, 2020. As summarized in Table A1 in the Appendix, partial lockdown includes “closed-off management” on highways, railways and public transport systems; and sets up checkpoints to control the inflow population, and implements surveillance and tighter controls in each neighborhood.

17 Our log-log specification is based on the classical susceptible-infectious-removed (SIR) model in epidemiology.

18 Date *t* = 23 indicates the date of January 23, 2020, and *t* = 60 the date of February 29, 2020.

19 Recall that outflows from Wuhan are not just residents of Wuhan; any travelers who entered Wuhan for whatever reason and then leave Wuhan would be included in the Wuhan outflows measured by Baidu Migration data.

20 See Panel C of Table 2 for the evidence from Wuhan lockdown on within-Wuhan population movement.

## References

Bajardi, P., C. Poletto, J. J. Ramasco, M. Tizzoni, V. Colizza, and A. Vespignani (2011). Human mobility networks, travel restrictions, and the global spread of 2009 h1n1 pandemic. PloS one 6 (1).

Chan, J. F.-W., S. Yuan, K.-H. Kok, K. K.-W. To, H. Chu, J. Yang, F. Xing, J. Liu, C. C.-Y. Yip, R. W.-S. Poon, et al. (2020). A familial cluster of pneumonia associated with the 2019 novel coronavirus indicating person-to-person transmission: a study of a family cluster. The Lancet 395 (10223), 514–523.

Charu, V., S. Zeger, J. Gog, O. N. Bjørnstad, S. Kissler, L. Simonsen, B. T. Grenfell, and C. Viboud (2017). Human mobility and the spatial transmission of influenza in the united states. PLoS computational biology 13 (2), e1005382.

Chinazzi, M., J. T. Davis, M. Ajelli, C. Gioannini, M. Litvinova, S. Merler, A. P. y Piontti, K. Mu, L. Rossi, K. Sun, et al. (2020). The effect of travel restrictions on the spread of the 2019 novel coronavirus (covid-19) outbreak. Science.

Ferguson, N. M., D. A. Cummings, C. Fraser, J. C. Cajka, P. C. Cooley, and D. S. Burke (2006). Strategies for mitigating an influenza pandemic. Nature 442 (7101), 448–452.

Gray, C. L. and V. Mueller (2012). Natural disasters and population mobility in bangladesh. Proceedings of the National Academy of Sciences 109 (16), 6000–6005.

Hollingsworth, T. D., N. M. Ferguson, and R. M. Anderson (2006). Will travel restrictions control the international spread of pandemic influenza? Nature medicine 12 (5), 497–499.

Huang, C., Y. Wang, X. Li, L. Ren, J. Zhao, Y. Hu, L. Zhang, G. Fan, J. Xu, X. Gu, et al. (2020). Clinical features of patients infected with 2019 novel coronavirus in wuhan, china. The Lancet 395 (10223), 497– 506.

Lai, S., N. W. Ruktanonchai, L. Zhou, O. Prosper, W. Luo, J. R. Floyd, A. Wesolowski, M. Santillana, C. Zhang, X. Du, H. Yu, and T. A. J (2020). Effect of non-pharmaceutical interventions for containing the covid-19 outbreak in china. doi: https://doi.org/10.1101/2020.03.03.20029843, medRxiv.

Lauer, S. A., K. H. Grantz, Q. Bi, F. K. Jones, Q. Zheng, H. R. Meredith, A. S. Azman, N. G. Reich, and J. Lessler (2020). The incubation period of coronavirus disease 2019 (covid-19) from publicly reported confirmed cases: Estimation and application. Annals of Internal Medicine.

Li, R., S. Pei, B. Chen, Y. Song, T. Zhang, W. Yang, and J. Shaman (2020). Substantial undocumented infection facilitates the rapid dissemination of novel coronavirus (sars-cov2). Science.

Liu, Y., A. A. Gayle, A. Wilder-Smith, and J. Rocklöv (2020). The reproductive number of covid-19 is higher compared to sars coronavirus. Journal of Travel Medicine.

Lu, X., L. Bengtsson, and P. Holme (2012). Predictability of population displacement after the 2010 haiti earthquake. Proceedings of the National Academy of Sciences 109 (29), 11576–11581.

Munshi, K. (2003). Networks in the modern economy: Mexican migrants in the us labor market. The Quarterly Journal of Economics 118 (2), 549–599.

Qiu, Y., X. Chen, and W. Shi (2020). Impacts of social and economic factors on the transmission of coronavirus disease (covid-19) in china. doi: https://doi.org/10.1101/2020.03.13.20035238, medRxiv.

Wang, Q. and J. E. Taylor (2016). Patterns and limitations of urban human mobility resilience under the influence of multiple types of natural disaster. PLoS one 11 (1).

WTO (2003). Consensus document on the epidemiology of severe acute respiratory syndrome (sars). Technical report, World Health Organization.

